# Quantifying the drivers of protective behaviours in epidemics: Mask wearing during COVID-19

**DOI:** 10.1101/2025.11.26.25341041

**Authors:** Marie Scantamburlo, Daniele Proverbio, Giulia Giordano

## Abstract

Epidemic dynamics are shaped by behavioural responses to the risk of contagion, which are in turn influenced by beliefs, disease awareness, compliance with government mandates, and other drivers. However, quantitative assessments of the importance of drivers of protective behaviours remain scarce, largely due to limited and fragmented datasets. This limitation hinders the development of empirically-grounded models that couple the dynamics of human behaviour and of epidemic spread, which would have the potential to substantially improve the prediction of disease transmission and the effectiveness of pharmaceutical and non-pharmaceutical interventions. Existing behavioural-epidemiological models typically rely on strong assumptions regarding the linear and direct dependence of behavioural responses on awareness, opinion dynamics, epidemic progression, or other factors that can only be partially validated against small-scale datasets. Here, we address this challenge by integrating multiple datasets to quantify the impact of different drivers of protective behaviours, focusing on mask-wearing during the COVID-19 pandemic. Combining multivariate regression, Granger causality and random forests for covariate analysis reveals that beliefs regarding the effectiveness of protective behaviours and compliance with government interventions explain most of the variability of mask-wearing behaviour worldwide and are substantially more predictive than peer influence or disease awareness. Our findings provide quantitative evidence on the primary drivers of mask wearing, establish a reproducible framework to identify the drivers of other protective behaviours, and empirically test key assumptions underlying widely adopted behavioural-epidemiological models, thereby supporting their refinement and interpretability.

**Plain language summary:** Behavioural responses during epidemics are shaped by attitudes, belief systems, cultural factors, opinions and disease awareness; however, the primary drivers of protective behaviours remain poorly understood due to limited comprehensive data and insufficient hypothesis testing. This limitation affects epidemic management as well as the development of epidemiological models that account for human behaviour, which are essential for improving predictions of disease spread and informing control strategies. By integrating multiple datasets collected during the COVID-19 pandemic and applying multivariate regression, Granger causality and random forests for covariate analysis on a carefully curated subset, we identify the key determinants of mask-wearing behaviour worldwide. Our findings improve the understanding and prediction of protective behaviour uptake and provide an empirical basis to test crucial hypotheses underlying behavioural-epidemiological models, and develop more accurate and effective modelling frameworks.

## 1 Introduction

Quantifying the impact of human behaviour on epidemic dynamics remains a major challenge in modern epidemiology. As observed during the COVID-19 pandemic, the progression of epidemics, as well as the effectiveness of pharmaceutical and non-pharmaceutical interventions, depends strongly on behavioural responses [1, 2, 3]. This observation has spurred the development of behavioural-epidemiological models, which aim to couple behavioural and epidemic dynamics to improve the understanding, prediction and control of infectious disease spread [4, 5], and in particular support the development of evidence-based mitigation interventions. Despite recent advances in the understanding of behavioural-disease dynamics and in the development of more advanced models [6], important limitations remain [7]. Incomplete data from heterogeneous and non-standardised sources provide little and non-unified evidence to behavioural scientists and modellers; this fact also hinders the development of purely data-driven models and leads to challenges in capturing nonlinear dynamics, avoiding overfitting, and ensuring interpretability and transferability across contexts and countries. Conversely, simpler mechanistic models often rely on strong assumptions that remain insufficiently validated against empirical data [8]; for instance, many dynamical models focus exclusively on opinions, awareness, or fear, assuming that each of these factors in isolation directly translates into behavioural changes that affect epidemic dynamics [6]. Recent attempts to integrate multiple behavioural dimensions within unified dynamical behavioural-epidemiological frameworks include game-theoretic approaches [9, 10], multi-agent models [11, 12], and compartmental models that explicitly incorporate adherence to protective measures [13, 14, 15] or mechanisms informed by behavioural science [16, 17]. However, the empirical validation of their underlying assumptions and predictive capabilities remains limited.

Bridging the gap between behavioural data and the assumptions underlying mathematical models is thus a crucial step to better understand the impact of drivers and determinants to adopt and support protective behaviours, as well as to develop evidence-based coupled social and epidemiological models. To support this aim, recent time-series datasets are particularly precious. In particular, the COVID-19 pandemic generated an unprecedented quantity of epidemiological, behavioural, and policy-related data [18]. Individual datasets have been used to model specific behavioural responses, including changes in mobility [19, 20], contacts [21] and economic activities [22, 1]. Still, substantial uncertainty persists regarding the relative importance and quantitative impact of the drivers of protective behaviours, including epidemic awareness, personal beliefs, trust in information sources, and public health interventions [7, 23]. In behavioural science, identifying the determinants of protective actions – that is, the psychological constructs hypothesised to influence protective behaviours [24] – is an active area of research; however, studies often rely on cross-sectional surveys administered only once to a cohort of participants (see [25] and references therein). Moreover, other potential drivers, such as government mandates or news from media outlets, may further shape behavioural responses [26, 6]. Integrating longitudinal and heterogeneous datasets would instead enable a quantitative assessment of the drivers of protective behaviours over time and across multiple dimensions. Combined with epidemiological and policy time-series data, such analyses would provide insight into behavioural-epidemic dynamics and enable empirical testing of modelling assumptions regarding the nonlinear interplay between behavioural, social and epidemic phenomena.

By analysing global datasets about mask-wearing protective behaviours, multiple candidate drivers of such behaviours, and epidemic evolution, we aim to quantify the impact and predictive capacity of the various drivers, identify the most relevant ones, and test common hypothesis used in behavioural-epidemiological models. We construct and systematically analyse a curated database, that integrates multiple public datasets containing worldwide information on the temporal evolution of (i) behavioural responses, (ii) behavioural determinants, including beliefs, awareness and trust in information sources, government interventions, including non-pharmaceutical countermeasures and testing policies and epidemic dynamics, during the COVID-19 pandemic. Focusing on mask wearing as a self-protective behaviour, we develop a comprehensive and comparable set of indicators to characterise the relationship between mask wearing and its potential drivers, identifying the most influential ones, employing interpretable univariate and multivariate analysis. Our results identify and rank the most important drivers of behaviours, highlight the nonlinear dependencies between them, and identify a minimal set of indicators that can help predict mask-wearing behaviours. Beyond identifying the main drivers of protective behaviour, our framework provides a reproducible approach to test behavioural-epidemic hypotheses, improve prediction of behaviour adoption, and develop more accurate and empirically grounded behavioural-epidemiological models.

## 2 Results

We first integrate multiple datasets containing information on different aspects of the COVID-19 pandemic over time, in different countries worldwide; details regarding each dataset, including access, coverage and limitations, are provided in Methods. We then analyse the relationship between mask-wearing behaviour and its potential drivers across countries and over time.

Previous studies on the determinants and drivers of behaviours have largely relied on surveys collected at only a few time points [27, 28] and on data analysed using univariate approaches such as regression or structural equation models. Few studies explicitly investigate long-term behavioural-epidemic temporal dynamics [3, 7] and mostly consider the effect of a single driver at a time [21, 29]. Importantly, determinants and drivers are known to overlap both conceptually and statistically in shaping behavioural responses [25], hence requiring multivariate approaches to disentangle and quantify the concurrent contributions of different factors. To address these challenges, we first assess the influence of each candidate driver on behavioural responses, to characterise their individual temporal relationships with mask wearing. We employ three progressively more informative approaches: (1) analysis of Pearson’s correlation coefficients *ρ*_*XY*_ , to test the hypothesis, common in the behavioural-epidemiological modelling [23], that mask wearing linearly correlates with their drivers; (2) multivariate linear regression (MLR), to model the joint linear dependence of mask-wearing behaviour on multiple drivers; and Granger causality analysis, to test whether time series of each driver improve predictions of mask-wearing time series. Although these methods explicitly account for temporal dynamics, they have limited ability to capture nonlinear interactions across heterogeneous classes of drivers. We therefore additionally employ an interpretable Random Forest machine learning method to identify and quantify nonlinear relationships, to identify the main drivers of behaviour adoption and rank their relative importance in reconstructing mask-wearing dynamics. Additional details about the employed approaches are provided in Methods.

### 2.1 Construction of the dataset

We integrate several global and public datasets to account for different epidemic factors and candidate drivers.

The main publicly available datasets providing global and densely sampled time series of maskwearing behaviour are the Delphi US CTIS [30], for the United States, and the Global COVID-19 Trends and Impact Survey (UMD-CTIS), for the rest of the world, developed by the Social Data Science Center at the University of Maryland in collaboration with Facebook [31]. These datasets provide daily-sampled time-series information on COVID-related behaviours, collected through questionnaires administered via Facebook between 2020 and 2022; owing to their temporal coverage, dense geographical granularity and sample size, they have been extensively used in behavioural research in recent years, see e.g. [32, 29]. Although the validity of these surveys for precise quantitative estimations (e.g., of vaccine uptake) has been questioned [33], our analysis focuses on temporal trends, which are more robust to scaling biases. Moreover, we employ analysis methods that are robust against random fluctuations in the data due to, e.g., daily samples or country-wise coverage, therefore providing useful results about the dynamical relationships between the variables of interest. For both datasets, we focus on the “mask wearing” behavioural indicator, which measures the percentage of respondents, in each country and for each day, reporting mask use all or most of the time in public places.

Based on recent surveys and studies [8, 6, 17, 34, 35, 25], we compile the set *X* of candidate drivers that are most commonly associated with protective behaviours in general, and mask wearing in particular, within the behavioural-epidemiological literature and models. We then systematically search public datasets for corresponding time series with sufficient temporal and population-level coverage. Among the psychological determinants discussed in the literature, only a limited subset is available as curated longitudinal data: from the CTIS datasets, we extract time-series indicators related to beliefs about mask effectiveness and about perceived risk and severity of COVID-19 infection, as well as awareness of the pandemic progression and trust in information sources. We further include epidemiological indicators, such as incidence and daily deaths associated with COVID-19, which are frequently modelled as direct drivers of behavioural responses and, thus, of epidemic dynamics [36]; these epidemiological indicators are sourced from the Our World in Data (OWID) and John Hopkins University Center for Systems Science and Engineering (JHU CSSE) datasets [37]. Finally, we consider government interventions, containment policies and mask mandates through the composite Oxford Containment and Health Index (CHI) from the Oxford COVID-19 Government Response Tracker (OxCGRT) dataset [38]. The CHI includes semi-continuous quantitative information about government stringency, as well as about facial covering mandates, capturing top-down interventions and public recommendations and quantifying their overall stringency at the population level. Details about the OxCGRT indexes and the CHI selection are in SI Sec. S1.

Overall, the selected indicators summarise individual, peer-to-peer and shared drivers and determinants that may influence the dynamics of behavioural responses, enabling the characterisation of the factors that mostly drive, on average, mask wearing across heterogeneous populations worldwide.

To select the countries to include in the final dataset, we apply a rigorous procedure based on systematic criteria related to temporal coverage, data continuity and statistical significance. Fig. 1 presents a summary of the country selection process. Details about each decision step (diamond checkpoints in Fig. 1) are in Methods, as well as SI Sec. S3. We analyse a time window extending from May 21th, 2021 to June 25th, 2022, which corresponds to the period over which all considered datasets exhibit significant and coherent overlap. Spanning more than one year, this interval ensures uniform data availability across all indicators and captures an important phase of the pandemic, including the spread of the Delta variant and the emergence of Omicron [39]. Starting from the 166 countries available in the CTIS datasets, the selection procedure yields a final dataset comprising 34 countries across Europe, Asia and the Americas, which are statistically and dynamically representative and comparable. In Fig. 2, these countries are shown and ranked according to their level of interventionism, quantified as the average CHI over the analysed time period. This ranking will be useful to interpret the results concerning the CHI as a driver; further details about the interventionism of the selected countries can also be found in SI Sec. S2.

**Figure 1.**
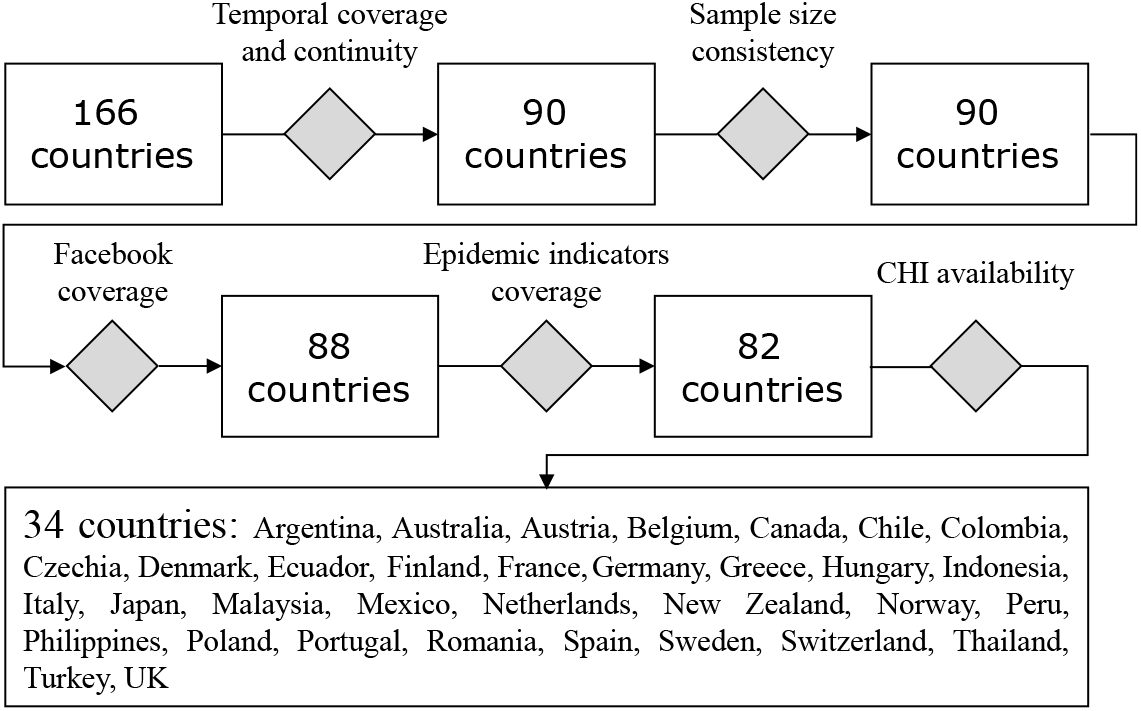
Flow diagram of the country selection process, which is described in detail in Methods and in SI Sec. S3.

**Figure 2.**
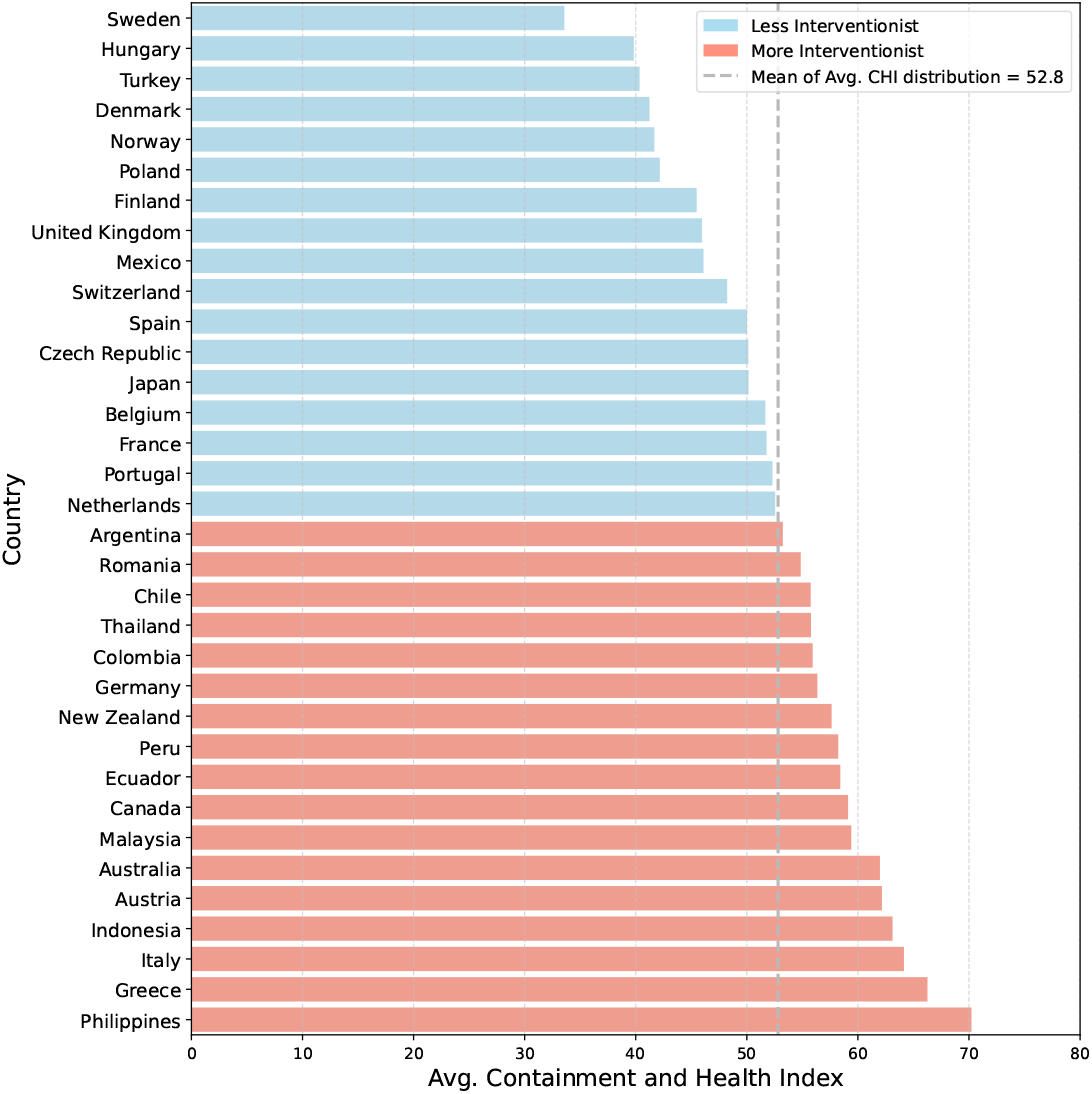
Ranking of countries according to the average Containment and Health Index over time: 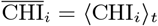 for country *i*. The vertical dashed line corresponds to the mean of the distribution across countries and serves as a visual reference for the level of interventionism of different countries. See SI Sec. S2 for further details.

### 2.2 Analysis of individual drivers’ classes

We first analyse the impact of individual classes of drivers (beliefs, awareness, trust, epidemiological indicators, interventions), using linear correlation and Granger causality analysis for each driver separately, and multivariate linear regression within each class of drivers. The results of the correlation analysis are summarised in Fig. 3, which visualises the distribution across countries of Pearson’s correlation coefficients *ρX*_*j*_ *Y* between driver *X*_*j*_ *∈ X* and mask-wearing behaviour *Y*. All results are clustered according to different classes of drivers, as discussed below.

**Figure 3.**
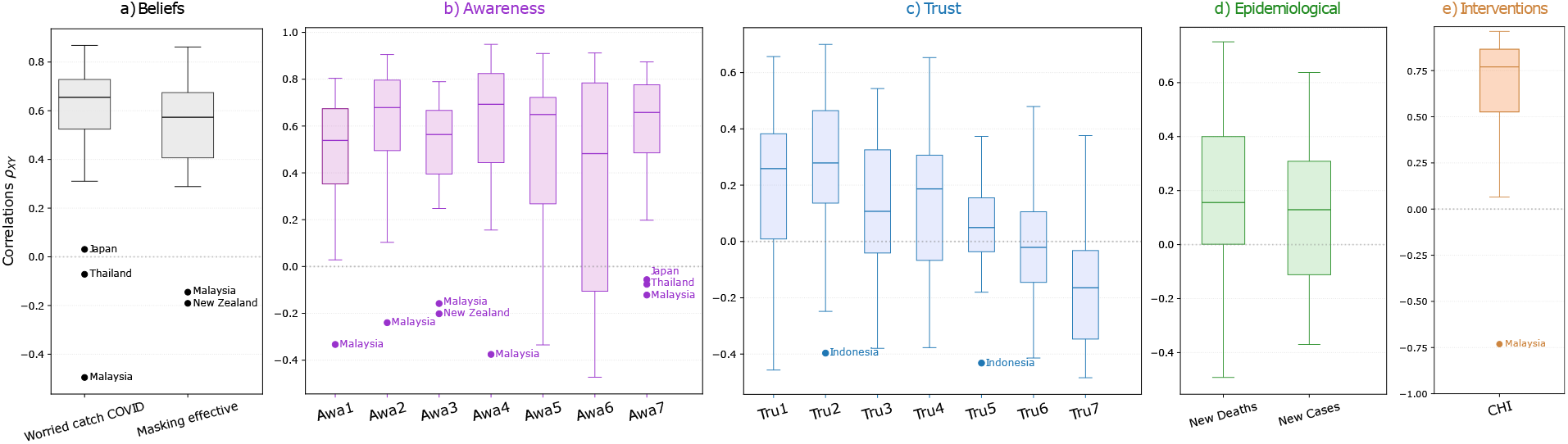
Boxplots of the distributions across countries of Pearson’s correlation coefficients *ρX*_*j*_ *Y* between maskwearing behaviour *Y* and each driver *X*_*j*_ .*∈ X* Drivers are clustered into classes, as described in the main text: a) Beliefs; b) Awareness; c) Trust; d) Epidemiological indicators; e) Government Interventions. Outlier countries are indicated below the boxplots.

#### Beliefs

The belief-related indicators capturing fear of COVID-19 infection and perceived effectiveness of mask wearing in reducing the risk of infection both exhibit relatively strong positive correlation with mask wearing, with median values around *ρ* = 0.6 and narrow whiskers (Fig. 3a). The outliers are Asian countries, where mask use is often habitual and therefore less directly associated with explicit beliefs, and New Zealand, where mask adoption increased sharply in September 2021 following government mandates (see SI Sec. S4A for a detailed discussion of outliers). Granger causality analysis confirms that beliefs Granger-cause the behaviour, but also reveals that mask-wearing behaviour Granger-causes both belief indicators (*p <* 0.05 in both cases), suggesting the presence of a feedback loop between beliefs and protective behaviour: the analysis suggests that the beliefs strongly support masks usage and, at the same time, mask usage strengthens the beliefs about fear of catching the disease and about masks effectiveness. This observation may be associated with social proofs and conformity [40], which have been also observed for other behaviours such as customers choices or vaccine uptake [41, 42]; further studies are needed to investigate the underlying psychological mechanisms. The observation that the belief about mask effectiveness greatly correlates with compliant protective behaviour is also in agreement with previous longitudinal studies [43] about the positive influence of efficacy beliefs for adherence. Multivariate linear regression (MLR) using the two belief indicators as predictors yields positive coefficients in both cases: perceived mask effectiveness exhibits a stronger linear relationship with behaviour (*β*_2_ = 1.35) than fear of infection (*β*_1_ = 0.32). Variance Inflation Factor (VIF) values, around 2.8 in both cases, indicate moderate collinearity, while the relatively high coefficient of determination *R*^2^ = 0.669 supports the hypothesis that beliefs represent an important driver of behavioural uptake.

#### Awareness

Awareness of the pandemic is assessed through information received from seven different sources (see Fig. 3b): local health workers (Awa1), experts (Awa2), WHO (Awa3), government health authorities (Awa4), politicians (Awa5), journalists (Awa6), friends and family (Awa7). Median correlations exceed *ρ* = 0.4 for all awareness indicators, suggesting a generally positive association with mask wearing. Most distributions are relatively compact, indicating moderate variability across countries; however, awareness mediated by politicians and journalists displays substantially broader distributions, including negative correlations, thus highlighting strong heterogeneity across countries (see SI Sec. S4B for further discussion). The identified outliers coincide with those observed for beliefs. Granger causality analysis reveals a predominantly unidirectional influence from information received through experts, WHO, and governmental institutions toward mask-wearing behaviour, whereas awareness mediated by local health workers, politicians, journalists, and friends and family exhibits bidirectional relationship with mask-wearing behaviour (*p* < 0.05). This distinction is intuitively plausible, as institutional actors are more likely to provide the same recommendations independent of population behaviour, while journalists and peers may also react to the behaviour of the population. Finally, MLR identifies the strongest positive associations for awareness through friends and family (*β*_7_ = 1.22) and local health workers (*β*_1_ = 1.15), followed by journalists (*β*_6_ = 0.86) and experts (*β*_2_ = 0.64). By contrast, awareness through WHO has a negligible coefficient (*β*_3_ = 0.07), while awareness through government authorities and politicians yields negative coefficients (*β*_4_ = −0.68 and *β*_5_ = −0.46). This phenomenon may arise from a suppressor effect, whereby variables that correlate positively with the outcome (in our case, mask wearing; *cf*. Fig. 3) have negative coefficients within multivariate models because they primarily explain other predictors rather than the dependent variable itself [44]. The coefficient of determination is *R*^2^ = 0.54, while VIF values, all below 5, suggest low-to-moderate multicollinearity, with the largest value (4.2) associated with awareness through local health workers. Although these values represent averages across countries with heterogeneous informational ecosystems (*cf*. SI Sec. S4B), they nonetheless provide insight into the relative effectiveness of different communication channels during pandemics, highlighting that sources perceived as closer to individuals may have a higher impact in influencing their behaviours.

#### Trust

In addition to awareness, we investigate whether trust in the same seven information sources influences mask-wearing behaviour. Overall, trust displays weak and highly heterogeneous correlations with behaviour: median values lie between 0.2 and −0.2, while the corresponding distributions are broad and dispersed, with large whiskers (Fig. 3c). Together, these results provide little evidence for a robust direct relationship between trust in information sources and mask wearing. Granger causality analysis identifies significant effects only for trust in politicians, journalists and health authorities; however, the corresponding MLR coefficients are negative. The value *R*^2^ = 0.28 suggests a low explanatory power of the regression and VIF values additionally indicate moderate-to-high multicollinearity, supporting the conclusion that trust alone is a weak driver of mask wearing. Similar conclusions are drawn when considering awareness weighted by trust, rather than trust alone: the influence is largely heterogeneous across countries and, in most of the cases, scarcely relevant (see SI Sec. S4C). While the literature on healthcare for individual patients has shown that trust in doctors and health systems plays a pivotal role [45], our results suggest that social proximity, more than the trust in a source, has a dominant influence on mask uptake.

#### Epidemiological indicators

We also investigate whether epidemiological indicators directly influence mask-wearing behaviour. Behavioural-epidemiological models frequently assume that infection incidence or prevalence, as well as the number of deaths, directly influence behavioural responses [6].

Moreover, during the COVID pandemic, information regarding new detected cases, deaths, and testing activity was disseminated daily through newspapers, websites, and public dashboards, and possibly influenced behavioural responses. We here focus on the effect of daily *New Cases* and daily *New Deaths*, both smoothed over a 7-day window to suppress fluctuations that may be associated with randomness or testing routines. The median correlations between both epidemiological indicators and mask wearing are positive, but weak (*ρ ≈* 0.2), suggesting that their relationship with behaviour is nonlinear or indirect. The corresponding distributions are also highly heterogeneous, see Fig. 3d: some countries exhibit strong positive correlations, while others display weak (approximately zero) or even negative correlations. This variability indicates that increases in case numbers or deaths do not necessarily translate systematically into stronger protective behaviour, nor decreases in disease incidence or mortality immediately translate into less frequent masking. Granger causality analysis reveals that daily new deaths unidirectionally Granger-cause mask wearing behaviour, whereas daily new cases exhibit bidirectional Granger causality with mask wearing; this result qualitatively supports previous modelling approaches that analysed a feedback between social responses and disease incidence [46], although the feedback explains only little of the behaviours observed in reality. In fact, MLR yields a very low explanatory power (*R*^2^ = 0.057) and the estimated model coefficients are extremely small(< 10^−4^), suggesting that the two epidemiological indicators explain only a limited fraction of the observed variability in mask wearing and that there is very little direct influence. These results suggest that relatively few individuals read and act upon epidemiological reports directly; the effect of epidemiological information appears to be likely mediated by informational and social processes shaping opinion dynamics, beliefs and perceptions.

#### Interventions

Finally, we analyse the role of the Containment and Health Index (CHI), which summarises government interventions, including mask mandates. The CHI captures the average “topdown” influence of institutional responses aimed at controlling the pandemic through recommendations, regulations, and enforcement measures ranging from fines to physical confinement [47, 48, 49, 50, 51, 52]. The median correlation of CHI with mask wearing is very high (*ρ* > 0.75), with relatively low variability across countries, indicating a strong and consistent positive linear relationship between government interventions and mask wearing in almost all countries. Malaysia is the only major outlier, exhibiting a strong negative correlation (*ρ* = − 0.73), which may reflect country-specific cultural, political or social dynamics that shaped negative responses to governmental directives [53]. For Malaysia, MLR returns *β* = −0.002, with *R*^2^ = 0.53, indicating a slight decrease in mask wearing as government interventions intensified. Conversely, all other countries exhibit positive coefficients (*β* = 0.013), with *R*^2^ = 0.34. Granger causality analysis on the entire dataset further confirms that CHI Granger-causes mask wearing behaviour unidirectionally. These results suggests that changes in government policies tend to precede and influence behaviour uptake: increases in policy stringency are typically followed by increases in mask usage, and relaxation of the stringency is usually followed by a decrease in the adoption of masks.

### 2.3 Multivariate Random Forest

The previous analyses, whether univariate or multivariate within individual class of drivers, provide insight into the isolated effects of different drivers on mask-wearing behaviour. However, the associated *R*^2^ values indicate that no single class of drivers alone is sufficient to fully explain behavioural dynamics. Moreover, as discussed in detail in SI Sec. S4C, several drivers exhibit mutual dependencies: for example, beliefs correlate significantly with CHI but weakly with epidemiological indicators, whereas awareness through journalists has very broad variability across countries. A fully multivariate analysis including all candidate drivers of mask wearing is therefore required to disentangle their concurrent contributions and identify the dominant determinants of behaviour. To this end, we employ a Random Forest (RF) approach (see Methods), using all candidate drivers (except trust-related ones, which already displayed negligible effects in the previous analysis) to reconstruct mask-wearing dynamics and identify the most influential drivers in predicting mask-wearing behaviour through the Variable Importance Vector (VIV).

The resulting ranking in shown in Fig. 4. By far the most influential drivers are the two belief-related indicators: perceived mask effectiveness and fear of infection. The third most influential driver is the CHI, highlighting the strong influence of population-wide government interventions on the adoption of protective behaviours; this results confirms empirical and independent analysis that suggest high correlation among containment scores and policy stringency [54]. Together, these three drivers explain 68% of the variability in behavioural dynamics. The remaining nine potential drivers contribute comparatively little to behaviour reconstruction, jointly accounting for the remaining 32% of the explained variance. Among them, awareness through socially proximal sources of information, such as local health workers and friends and family, is more relevant than information originating from institutional, socially distant sources (journalists, government health authorities, experts, WHO, politicians, in decreasing order of importance). Effective communication, grounded in principles of coherence and clarity during a social crisis, is known to foster adherence to public policies [55]; proximal sources of information may promote such communication. Finally, epidemiological indicators display relatively limited importance: the impact of new deaths is comparable to that of awareness through friends and family, while the impact of new cases is even lower and is comparable to that of awareness through journalists.

**Figure 4.**
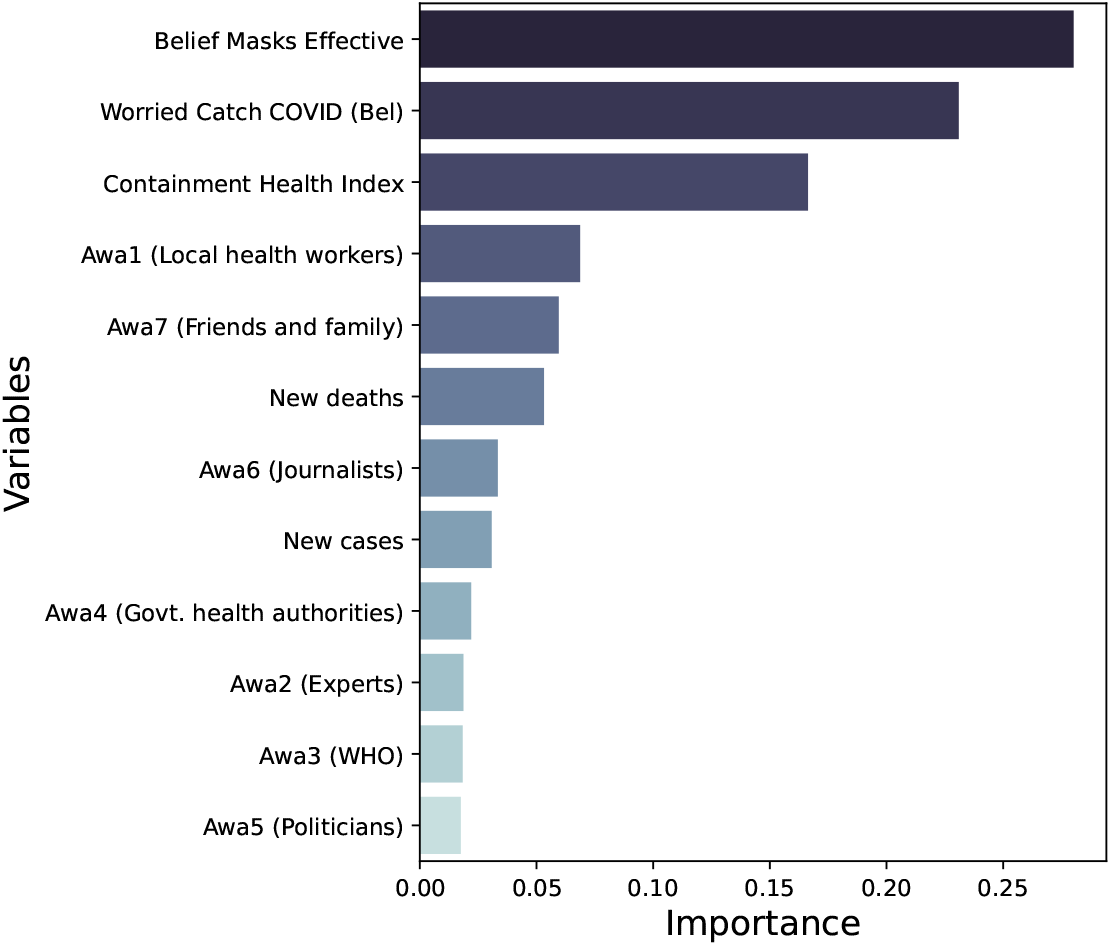
Ranking of the importance of different drivers in reconstructing mask-wearing behaviour, quantified through the Variable Importance Vector (VIV) obtained from the Random Forest (RF) model.

The RF model reconstructs mask-wearing dynamics with high accuracy and limited evidence of overfitting (*R*^2^ = 0.974 on the training set and *R*^2^ = 0.950 on the test set, OOB score of 0.955); the mean squared error (MSE) is also low (0.0020 on the training set and 0.0039 on the test set, with Bias^2^ equal to 0.0038 and variance 0.0001), confirming the robustness and accuracy of the reconstruction, which is effective in reproducing real behavioural trends. Fig. 5a compares empirical mask-wearing dynamics with their RF reconstructions for a representative subset of countries selected from Fig. 2 across different levels of interventionism. The reconstruction is overall very good, with an average *R*^2^ = 0.85 across the displayed countries. Notably, the reconstruction of mask-wearing dynamics through the RF model using as input only the three dominant drivers identified in Fig. 4, namely the two beliefs indicators and the CHI, still produces accurate results. As shown in Fig. 5b, considering only these three drivers helps predict mask wearing in various countries with high accuracy (average *R*^2^ = 0.78 over the considered countries), with only a slight underestimation of peaks. This result supports the identification of beliefs and stringency as the main drivers of mask wearing and provides an indication for future studies, suggesting which proxy variables can be monitored to assess the uptake of protective behaviours in a population.

**Figure 5.**
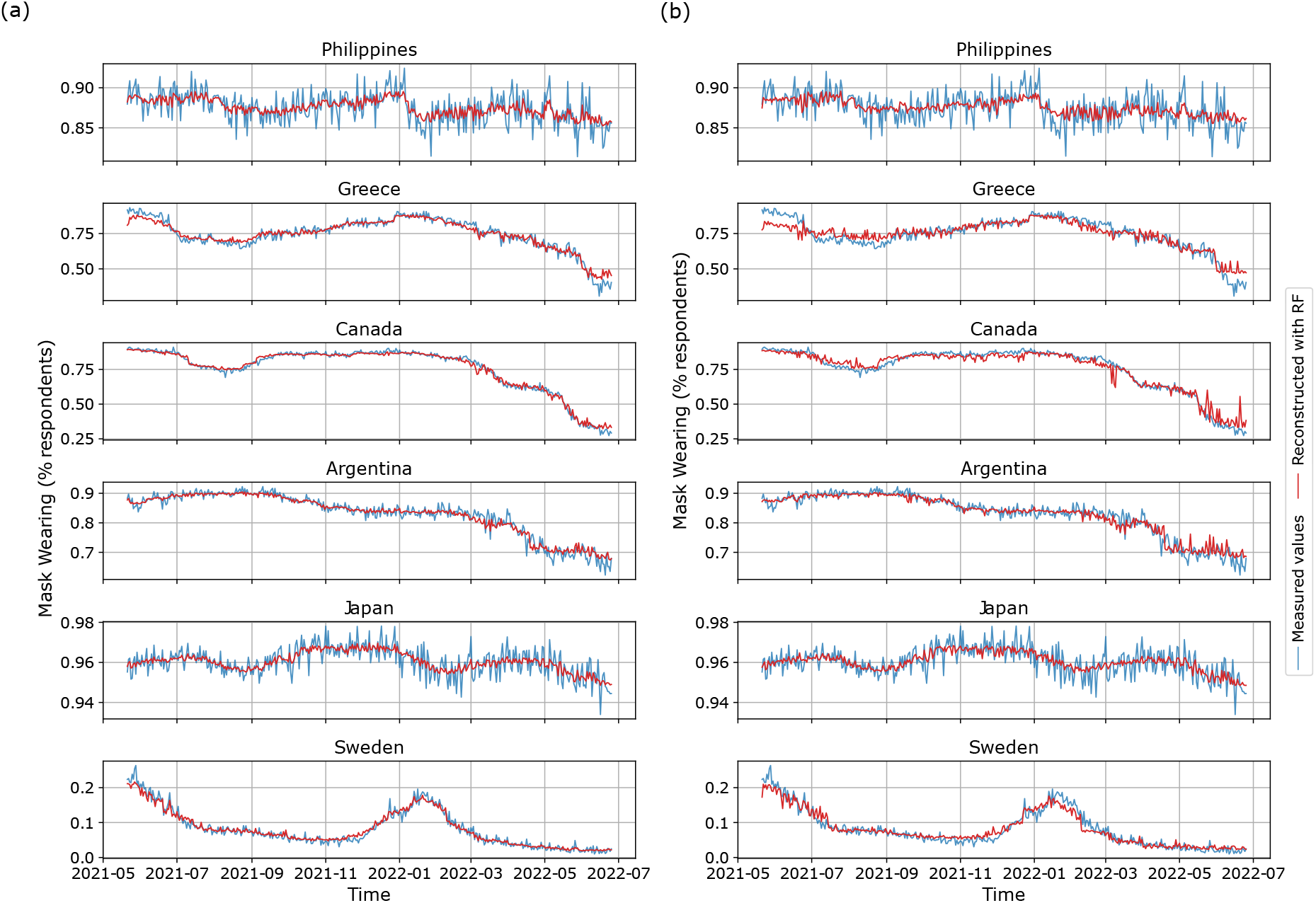
Time evolution of mask-wearing behaviour (blue) compared with its RF reconstruction (red) for the Philippines, Greece, Canada, Argentina, Japan, Sweden. (a) Using all drivers as features of the RF reconstruction; (b) using only the top three drivers (two beliefs indicators and the CHI) to inform the RF reconstruction.

## 3 Discussion

We have constructed a comprehensive and statistically robust dataset integrating behavioural, epidemiological, and policy-related information and performed a data-driven analysis of the principal drivers of protective behaviours during the COVID-19 pandemic, focusing on mask wearing. The combination of univariate and multivariate analysis identified belief-related determinants – in particular, perceived mask effectiveness and fear of infection – and government interventions as the dominant factors driving maskwearing dynamics. Conversely, awareness, trust and epidemiological indicators exhibit comparatively limited explanatory power. This ranking of drivers highlights primary areas of intervention to improve adoption of protective behaviours in future pandemics and can support the development of more accurate and empirically grounded behavioural-epidemiological models, focusing on the drivers most strongly associated with behavioural uptake.

Our combined analysis additionally helps interpret the role of drivers across countries. Some drivers display relatively consistent effects across countries (such as belief in mask efficacy, whose Pearson correlation coefficient distribution has much lower variability compared with the CHI), which supports their consideration for models and interventions with broad validity. Other drivers instead (such as awareness through journalists) show substantial heterogeneity across countries and therefore likely require country-specific interpretation, modelling and interventions, complementing previous questionnaire-based conclusions regarding social dynamics across different countries [26]. Hence, our findings may support the development of improved modelling approaches and public-health decision making at both qualitative and quantitative levels. For instance, from a policy perspective, our results suggest that interventions targeting beliefs (e.g., through nudges or behavioural incentives) may be more effective in promoting the adoption of protective behaviours than simply increasing the stringency of restrictions and mandates. Similarly, health communication and information campaigns may benefit from prioritising the conversion of awareness into an actual belief, in a funnel perspective [56], and from relying on socially proximal actors (such as local health workers or peers) rather than exclusively on institutional communication channels.

Our analysis also unravels substantial collinearity and mediated effects among the different behavioural drivers, as well as their nonlinear relationship with the protective (mask-wearing) behaviour. In fact, even the stronger predictors do not exhibit perfectly linear relationships with mask wearing, suggesting that their use as direct proxies for behaviour in behavioural-epidemiological models should be regarded only as a first approximation. Awareness, for instance, is extensively used in dynamical models [6], yet our results suggest that it constitutes only a weak proxy variable for explaining mask-wearing dynamics. More generally, individual beliefs and institutional interventions appear to exert stronger effects than peer pressure or peer-mediated awareness: this observation should be considered when developing empirically consistent behavioural-epidemic models. These results further emphasise the importance of validating models directly against behavioural data and of considering the original behavioural variable rather than surrogate variables, which likely introduce systematic biases. More broadly, our findings support the development of modelling approaches that integrate behavioural-science frameworks, such as the Health Belief Model, within epidemic dynamics [17], but highlight the need to incorporate nonlinear feedback mechanisms linking government interventions, behavioural responses, and epidemic dynamics, so as to capture the mediating role of beliefs and other drivers of behaviour. Determining the precise functional forms of these interactions is an important direction for future work.

Our Random Forest analysis also demonstrates that accurate reconstruction of behavioural dynamics can be achieved primarily from belief-related variables and government interventions. Future studies may build on these findings to improve behavioural forecasting during epidemics, for instance by identifying suitable and easily measurable proxies for beliefs, e.g. through social media analysis [57], and integrating them with intervention-related indicators, such as the CHI, within predictive machine-learning frameworks.

We acknowledge that the criteria adopted to construct the considered dataset have been intentionally stringent, since our objective was to jointly analyse all candidate drivers within a coherent and comparable framework. Future studies focusing on more restricted subsets of drivers may therefore extend the analysis to a larger number of countries, while following the same methodological approach. Moreover, although we have focused on the main determinants and drivers of behaviours commonly discussed in the behavioural-epidemiological literature and included in models, several additional factors may influence the adoption of protective behaviours. For instance, vaccination status may affect willingness to wear masks [58, 59] by modifying individual perceptions of infection risk. Economic conditions may also exert bidirectional effects [60]: lower-income individuals may wear masks more frequently if alternative protective strategies such as remote work or social distancing are inaccessible or too expensive, and economic constraints may also reduce mask wearing because of the perceived cost of protective equipment. Additional sub-determinants [25] may further contribute to behavioural variability at the individual level; as new longitudinal datasets become available, integrating these dimensions into behavioural-epidemiological analyses will help further clarify the complex interplay between epidemics and human behaviour.

## 4 Methods

### 4.1 UMD-CTIS and Delphi US CTIS Datasets

These datasets were developed within the “Global COVID-19 Trends and Impact Survey” initiative of the University of Maryland Social Data Science Center, in partnership with Facebook (Meta). They comprise more than 100, 000 daily questionnaire responses collected from Facebook users between 2020 and 2022. The UMD-CTIS dataset covers 115 countries worldwide [31] and is publicly accessible via API [61]. The Delphi US CTIS dataset provides analogous information for US states [30] and is accessible using the *epidatpy* Python package (https://cmu-delphi.github.io/epidatpy/). The datasets consist of time series of indicators, grouped into thematic domains such as symptoms, behaviours, and trust. For each indicator, the time series provide the proportion of respondents who answered positively to the associated question. The full list of indicators and their explanation is available at the project website (gisumd.github.io/COVID-19-API-Documentation/), which also discusses known limitations (such as coverage), anonymisation, weighting, curation and pooling protocols; see also [62]. We use the “daily” aggregation method to preserve temporal resolution and facilitate integration with different datasets, and we use pooled data for each considered country, without regional breakdown. The analysed indicators (expressed in percentages) along with their category are: *mask* (behaviour: respondents reporting wearing a mask all or most of the time in public); *worried catch COVID* (belief: respondents reporting strong or moderate concern about contracting COVID-19); *belief masking effective* (belief: respondents considering face masks very or moderately effective in preventing COVID-19 transmission); *received news* (*s*) (awareness: respondents reporting having received COVID-19-related information in the past 7 days from source *s*) and *trust COVID info* (*s*) (trust: respondents reporting trust in COVID-19-related information from source *s*), where *s* is one of the following: local health workers, experts, WHO, government health authorities, politicians, journalists, friends and family.

### 4.2 OWID and JHU CSSE dataset

The Our World in Data (OWID) project integrates epidemiological information from institutional sources worldwide, including WHO and national public-health agencies, into a unified public database [37] accessible via GitHub (https://github.com/owid/covid-19-data/blob/master/public/data/owid-covid-data.csv). From the OWID database, we extract time series describing daily incidence, deaths, number of tests performed and test positivity rate. Testing data for the United States are instead obtained from the Center for Systems Science and Engineering (CSSE) at Johns Hopkins University (https://github.com/govex/COVID-19/tree/master/data_tables/testing_data), which aggregates information from state-level public dashboards and records from the US Department of Health & Human Services.

### 4.3 OxCGRT dataset

The Oxford COVID-19 Government Response Tracker (OxCGRT) dataset [38], publicly accessible through the GitHub repository (https://github.com/OxCGRT/covid-policy-dataset/blob/main/data/OxCGRT_compact_national_v1.csv), provides time series of indicators quantifying government interventions adopted worldwide in response to the COVID-19 pandemic; see [63] and SI Sec. S1 for details. Among the available indicators (Facial Covering, Stringency Index, Containment and Health Index, and Government Intervention Index), this work focuses on the Containment and Health Index (CHI), which summarises containment policies and public-health measures including mask mandates.

### 4.4 Criteria for dataset construction

The analysed dataset is constructed by filtering the countries of interest through a few selection criteria, so as to improve the significance and comparability of the results; see the summarising flowchart in Fig. 1.

The first selection criterion concerns the temporal coverage and continuity of the CTIS datasets. Among all surveyed countries, we retain only those with data available for at least 300 days (approximately 75% of the analysed period) and with daily sampling, to ensure broad and consistent temporal coverage. To guarantee data continuity, countries exhibiting interruptions in data reporting longer than 7 consecutive days are excluded, thereby reducing the likelihood that missing values reflect systematic discontinuities in data collection. We further verify that the sample size and variability of the CTIS indicators are sufficiently well distributed and internally coherent within each country to support statistical reliability. As discussed in SI Sec. S3A, no country displaying anomalous or systematically inconsistent indicators was identified, further supporting the suitability of the CTIS datasets for time-series analysis.

Next, we assess the representativeness of Facebook-derived survey data, using the DataReportal (https://datareportal.com/social-media-users) and NapoleonCat (https://stats.napoleoncat.com/) services, both accessed in October 2025, to estimate Facebook coverage and diffusion of mobile devices in each country during 2021 and 2022. In fact, although the CTIS datasets are innovative and potentially broad in coverage [31], they may be limited by sample representativeness, since Facebook usage is not uniform across countries owing to cultural factors, digital divides effects, unequal device accessibility, and political or regulatory constraints [64, 65, 66]. Consequently, in countries where Facebook coverage is low, survey responses may not adequately represent the broader population. Countries with Facebook coverage below 50% of the population are therefore excluded, to reduce potential biases. Additional discussion regarding the use of Facebook-derived data is provided in SI Sec. S3B.

Additional filters are applied to ensure the reliability of epidemiological indicators. To verify that pandemic indicators adequately capture COVID-19 transmission, rather than reflecting insufficient testing activity, we exclude countries that never reported testing data, exhibited insufficient testing continuity or frequency, or contained excessive temporal gaps in records (see SI Sec. S3C). We further exclude countries for which the test positivity rate exceeded 15% during most of the analysed period, since such values may indicate substantial under-testing and inadequate epidemic surveillance according to WHO guidelines.

Finally, we only retain countries for which the OxCGRT CHI indicator is available at the state level. Since state-level CHI data are unavailable for individual US states during the analysed period, as stated in its Github documentation (*cf*. SI Sec. S3D), they are excluded from the final dataset.

Additional details on selection criteria (Fig. 1), detailed analysis and statistical tests are provided in SI Sec. S3.

### 4.5 Correlation analysis

To quantify linear correlations and test the common modelling assumption that drivers correlate linearly with behaviours [23], we compute the Pearson correlation coefficients 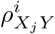 between mask wearing (*Y*) and each driver *X*_*j*_ *∈ X* , for each country *i*: 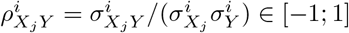. For each pair *X*_*j*_ *∈ X* and *Y* , we then obtain a distribution of values across all countries, visualised through the boxplots shown in Fig. 3. Outlier countries are discussed in detail as described in the Main Text.

### 4.6 Multivariate linear regression (MLR)

To go beyond pairwise correlations, we employ MLR to quantify the extent to which mask-wearing behaviour *Y* is jointly explained by multiple candidate drivers 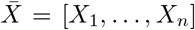 , seen as independent input variables [67]. MLR assumes a linear relationship of the form

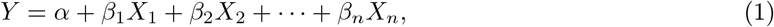

and estimates from data the values of the intercept *α* and of the regression coefficients *β*_1_, … , *β*_*n*_. Each coefficient *β*_*j*_ quantifies the expected variation in *Y* associated with a unit variation in *X*_*j*_, when all other variables are held constant. The model is only valid if all the variables *X*_1_, … , *X*_*n*_ are independent; we thus assess the risk of multicollinearity by computing the Variance Inflation Factor (VIF) [68]. We perform MLR and VIF analyses separately for each class of drivers (beliefs, awareness, trust, epidemiological indicators, interventions) using the Python sklearn package, which also provides the coefficient of determination *R*^2^ *∈* [0; 1] to quantify the fraction of behavioural variability explained by the MLR model. From the MLR output, we report in the main text the best-fit values of *β*_*j*_, whose associated relative standard deviations are below 1%.

### 4.7 Granger causality

The previous analysis characterises correlations and linear dependencies, but does not provide information about temporal predictability between variables. We employ Granger causality analysis to investigate direct temporal relationships between time series and, in particular, determine how well past values of *x*_*t*_ can predict future values of *y*_*t*_ [69]. Let *ℋ* denote the history of all the variables up to time *t −*1.

The quantity *P*(*y*_*t*_ |*ℋ*) then represents the optimal prediction of *y*_*t*_ conditioned on the complete history *ℋ*_*t*_, whereas*ℋ*_*t*_ *x* denotes the same history obtained by excluding past observations of *x*_*t*_, and hence exclusively based on past observations of *y*_*t*_. Then, *x*_t_ Granger-causes *y*_*t*_ if:

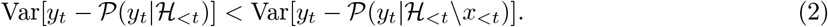

Intuitively, this means that including the past values of *x*_*t*_ improves the prediction of future values of *y*_*t*_. Statistical significance is assessed through hypothesis testing: the null hypothesis (assessed with p-value *>* 0.05) states that past values of *x*_*t*_ do not improve the prediction of *y*_*t*_ (i.e., they do not provide relevant additional information); rejection of the null hypothesis therefore indicates statistically significant Granger causality. To perform the analysis, we use time series data of all *X*_*j*_ *∈ X* and *Y*. To capture delays associated with behavioural and epidemiological processes, we consider a lag *t* up to 12 days, consistent with typical timespan (10–15 days) between contagion and death in severe COVID-19 infection [70]. Granger causality tests are implemented using the Python `statsmodels` library. For each pair of variables, all admissible lags are evaluated and the minimum p-value across lags is retained.

### 4.8 Random Forest

A Random Forest (RF) is a supervised machine-learning algorithm that combines the predictions of randomly constructed decision trees, each trained on a bootstrap sample of the training data [71]. Unlike linear regression approaches, RF does not assume linear relationships among variables and is comparatively robust to multicollinearity effects. RF models also show excellent performance when the number of variables is much larger than the number of observations [72]. Additionally, RF models are interpretable and allow quantification of the relative contribution of each predictor to the reconstructed output, through the Variable Importance Vector, VIV [71]. We use RF to identify which subsets of behavioural drivers can best reconstruct mask-wearing dynamics and to quantify their relative importance through the VIV. We assess the accuracy of the RF reconstruction using the coefficient of determination *R*^2^ *∈* [0; 1], the mean squared error (MSE), decomposed into Variance and Bias^2^ (i.e., the square of the difference between the true target data and the forest’s average prediction), and the OOB (Out-of-Bag) score, indicating better accuracy if close to 1. All metrics are computed using the `mlxtend` library [73]. We implement the RF model through the `RandomForestRegressor` function from the `ensemble` module of the `scikit-learn` Python package. We employ an 80:20 train-test split and use 300 decision trees. The model hyperparameters are selected using `scikit-learn`’s RandomizedSearchCV function to perform a Random Search 5-fold Cross-Validation, which yields *max depth*=12; *max features*=log2; *min samples leaf* =6.

## Data Availability

The composed database used for the analysis can be found at the link provided. All original datasets are available as disclosed.

https://github.com/daniele-proverbio/EpiBeha_analysis

## Code and data availability

The code and the composed dataset used for the analysis are available at https://github.com/daniele-proverbio/EpiBeha_analysis.

## Acknowledgments

We thank Matti T.J. Heino for useful discussion.

## Funding

Work supported by the European Union through the ERC INSPIRE grant (project number 101076926). Views and opinions expressed are however those of the authors only and do not necessarily reflect those of the European Union or the European Research Council Executive Agency. Neither the European Union nor the European Research Council Executive Agency can be held responsible for them.

## Author contributions

**MS:** Methodology, Software, Validation, Formal analysis, Investigation, Data Curation, Writing - Review & editing, Visualization. **DP:** Conceptualization, Methodology, Software, Validation, Investigation, Writing - Original draft, Writing - Review & editing, Visualization, Supervision. **GG:** Conceptualization, Methodology, Writing - Review & editing, Supervision, Project administration, Funding acquisition.

## Competing interests

The authors declare no competing interests.

## Notes

### Competing Interest Statement

The authors have declared no competing interest.

### Author Declarations

We used publicly available data from various online datasets. The UMD-CTIS dataset can be access via its page gisumd.github.io/COVID-19-API-Documentation/. Delphi US CTIS is accessible using the "epidatpy" Python package (https://cmu-delphi.github.io/epidatpy/). OWID epidemiological data can be accessed from https://github.com/owid/covid-19-data/blob/master/public/data/owid-covid-data.csv. Testing data about US are instead sourced from the Center for Systems Science and Engineering (CSSE) at Johns Hopkins University (https://github.com/govex/COVID-19/tree/master/data_tables/testing_data). The interventions dataset is publicly accessible on the OxCGRT GitHub repository (https://github.com/OxCGRT/covid-policy-dataset/blob/main/data/OxCGRT_compact_national_v1.csv)

### Summary of Updates

Additional analysis on the reconstruction of behavioural time series using only the three top features from the classifier.

