## Supplementary Information for "Quantifying the drivers of protective behaviours in epidemics: Mask wearing during COVID-19"

#### S1 OxCGRT intervention indexes

A central requirement for this study is to quantify the timing and intensity of government interventions related to mask wearing in each country. The Oxford COVID-19 Government Response Tracker (OxCGRT) [1] quantifies the evolution of the stringency of government interventions adopted over time in numerous countries worldwide to mitigate the COVID-19 pandemic. It includes several indexes summarising different aspects of government responses, as reported in Table S1, which lists the public interventions and policy measures considered by the Oxford group, and in Table S2, showing which policy components are included in each of the OxCGRT summary index.

The **Face Covering** measure, H6 in Table S2, is an ordinal indicator designed to quantify the extent to which mask use was required, or recommended, by governments. It is defined on an integer scale from 0 to 4, with each value corresponding to a specific level of policy stringency [2]; the full description of each level is provided in Table S3. Although useful for basic comparisons between countries, its five-level scale is relatively coarse and cannot capture the complexity of the regulatory framework adopted during COVID-19, as it provides only step-wise changes rather than semi-continuous dynamics.

The **Stringency Index**, ranging from 0 to 100, summarises, into a single numerical value that quantifies the severity of restrictions, containment and closure policies (C1 to C8) together with public information campaign policies (H1), without including mask use (and also excluding most of health-related policies and all economic policies). Hence, although widely used in the literature, it is not suitable for our purposes.

The **Containment and Health Index** (CHI), also ranging from 0 to 100, includes all the components of the Stringency Index, together with testing policy (H2), contact tracing (H3), facial coverings (H6), vaccination policy (H7) and protection of elderly people (H8).

The **Government Intervention Index**, ranging from 0 to 100, extends the CHI by additionally including economic measures (E1 and E2).

Of all the reported indexes, our analysis uses the Containment and Health Index (CHI), because it includes facial covering policies (H6) while avoiding the additional complexity introduced by the economic indexes E1 and E2.

Figures S1 and S2 compare the Stringency Index, Containment and Health Index, Government Intervention Index (all semi-continuous, with a range from 0 to 100) and the Face Covering measure quantification (integer-valued from 0 to 4) over time for each country. The indexes display richer dynamics and capturing multiple interventions that may, directly or indirectly, influence mask-wearing behaviour; examples include public-transport policies requiring mask use on buses or trains, and public information campaigns promoting prevention. Since these indexes summarise multiple policies, they also show substantial variability over time and across countries. They are therefore useful for characterising the overall regulatory context and for analysing how changes in government responses influenced pandemic dynamics over time, across national settings.

The three composite indexes display similar trends in most countries, although the Stringency Index does not include the facial covering indicator H6 (which limits its usefulness for the objectives of this study) and the Government Intervention Index includes additional economic components while exhibiting a temporal profile that is nearly identical to that of the Containment and Health Index (CHI), apart from small offsets in some countries, such as Japan.

Hence, the CHI used for our analysis is sufficiently representative of government interventions related to protective measures at large. As a side observation, economic interventions appear to have limited additional impact on the temporal structure of the intervention stringency in this context and are therefore neglected as a first approximation, although they may be investigated in future work.

Index data are from the Blavatnik School of Government, University of Oxford (2023), with minor processing by Our World in Data: “COVID-19 Containment and Health Index” [OWiD dataset]; Blavatnik School of Government, University of Oxford, “Government Response Tracker (OxCGRT)” [original data]. Data were retrieved as of May 2025 from https://archive.ourworldindata.org/20250909-093708/grapher/covid-containment-and-health-index.html (archived on September 9, 2025). We also retrieved data from the official OxCGRT GitHub repository (https://github.com/OxCGRT/covid-policy-dataset/tree/main). The documentation for US states (https://github.com/OxCGRT/USA-covid-policy) reports “Currently we provide coding up to 31 July 2020 for 13 indicators: C1 through C8, E1, E2, and H1 through H3.” Because H6, corresponding to facial coverings, is not available at the US state level for the period considered in this study, US states cannot be reliably included in our analysis.

#### S2 Ranking of countries by government interventions

To characterise the level of interventionism across the analysed countries, we rank them according to their CHI values. For each country, we compute the temporal average, minimum, and maximum of the CHI over the analysed period, and use these quantities to rank countries from most to least interventionist.

For visual reference, we introduce a threshold distinguishing “more” and “less” interventionist countries. This threshold is defined as the overall mean of the distribution. In the case of the distribution of average CHI values, the mean is 52.8. The distributions of national averages, minima, and maxima follow a normal distribution, as we have verified using Q-Q plots and Shapiro-Wilk tests; see Figures S3 for average values, and S4 for minimum and maximum values. Minimum CHI values are mostly below the threshold of 52.8, while maximum CHI values are all above it. Moreover, the distribution of maximum values appears flatter and with thinner tails than that of minimum values; in fact, the kurtosis of the distribution of maxima is lower than that of the distribution of minima. Figure S5 shows the ranking of countries according to the average CHI, while Figure S6 shows the rankings according to the minimum and maximum CHI.

Importantly, a country’s position in this ranking does not quantify the effectiveness of the adopted interventions; it just summarises the stringency of the governmental response to the pandemic. As shown in Figure S5, the Philippines ranks highest in terms of mean intervention level; its maximum CHI exceeds 80 out of 100 (see Figure S2) and reflects the implementation of one of the longest and most stringent lockdowns (https://www.wider.unu.edu/publication/dutertes-pandemic-populism) and school closures [3] worldwide. During the pandemic, the Philippine government also adopted highly restrictive mask policies, making masks mandatory in all situations [4]; accordingly, its Face Covering level reaches the maximum value of 4 (see Figure S2).

By contrast, Sweden adopted a less interventionist approach than most other countries considered in this study. The Swedish government did not impose strict lockdown measures, relying instead on voluntary recommendations such as physical distancing, remote working, and bans on public events, while kindergartens and schools were kept open throughout the pandemic [5]. Sweden required face coverings only in specific situations [5]; consequently, its maximum Face Covering value is 2 (see Figure S2).

Comparing Figures S5 and S6 helps interpret the stringency of intervention adopted by individual countries. For example, Turkey reached a maximum CHI value of approximately 80%: in fact, at the end of April 2021, the country declared a full lockdown and suspended educational activities [6]; after 20 days, it began a gradual normalisation process, during which the government introduced a curfew and later reopened food services [6]. This pattern is visible in Figure S2: from July 2021 onward, the CHI decreases substantially, reaching a minimum of approximately 25%, as also shown in Figure S6. Conversely, Italy maintained a consistently high level of intervention between May 2021 and June 2022 [7, 8]. The government adopted the Green Pass to allow easier access to public spaces for vaccinated individuals or those with a recent negative COVID-19 test [9, 10]. Schools reopened with mandatory masks and interpersonal distancing requirements (https://www.gazzettaufficiale.it/eli/id/2021/). Italy also continued to require face coverings in most indoor public spaces and on public transport, as reflected in Figure S1. Given the intensity and persistence of these measures, Italy appears among the countries with the highest minimum and maximum CHI values.

### S3 Criteria for database construction

As described in the main text, the first filtering criterion used to construct the final database is based on temporal coverage and data continuity. Specifically, we retain only countries with data available for at least 300 days and with no interruptions in data reporting longer than 7 consecutive days. After this first filtering step, the retained countries and US states are: Argentina, Australia, Austria, Belgium, Brazil, Canada, Chile, Colombia, Czech Republic, Denmark, Ecuador, Egypt, Finland, France, Germany, Greece, Hungary, India, Indonesia, Italy, Japan, Malaysia, Mexico, Netherlands, New Zealand, Norway, Peru, Philippines, Poland, Portugal, Romania, Spain, Sweden, Switzerland, Taiwan, Thailand, Turkey, Ukraine, United Kingdom, Venezuela, Vietnam, Alaska, Alabama, Arkansas, Arizona, California, Colorado, Connecticut, Delaware, Florida, Georgia, Hawaii, Iowa, Idaho, Illinois, Indiana, Kansas, Kentucky, Louisiana, Massachusetts, Maryland, Maine, Michigan, Minnesota, Missouri, Mississippi, Montana, North Carolina, North Dakota, Nebraska, New Hampshire, New Jersey, New Mexico, Nevada, New York, Ohio, Oklahoma, Oregon, Pennsylvania, Rhode Island, South Carolina, South Dakota, Tennessee, Texas, Utah, Virginia, Vermont, Washington, Wisconsin, West Virginia.

The additional selection criteria are detailed in the subsections below.

#### S3.1 Sample size consistency

Since the UMD Global CTIS survey includes questions associated with different indicators [11], the number of respondents varies across questions, countries and time. Consequently, sample size depends on the country, indicator, and survey date. To ensure that the dataset provides a sufficiently reliable representation of population-level behaviour, we assess the statistical significance of sample sizes. Countries with sample sizes that are too small, or with high internal variability across indicators, may provide non-representative data and are therefore excluded from the analysis. We first verify that sample sizes are sufficiently large to be statistically significant, and then assess the internal coherence of sample-size values within each country.

For each country ($c \in C$) and indicator ($j \in J$), we compute the mean sample size $\bar{s}_{c,j}$ over all time steps $t = 1, \ldots, n_{c,j}$ as

$$\bar{s}_{c,j} = \frac{1}{n_{c,j}} \sum_{t=1}^{n_{c,j}} s_{c,j}(t). \tag{S1}$$

For each country, we then compare the average sample size of the mask indicator with the average sample size of the other indicators using the absolute differences:

$$d_{c,j} = |\bar{s}_{c,j} - \bar{s}_{c,\text{mask}}|, \quad j \neq \text{mask}. \tag{S2}$$

Averaging all these absolute differences across all non-mask indicators yields the average internal dispersion of mean sample sizes for each country:

$$D_c = \frac{1}{|J| - 1} \sum_{\substack{j \in J \\ j \neq \text{mask}}} d_{c,j}, \tag{S3}$$

where $J$ is the set indexing all possible indicators and $|J|$ is its cardinality. The distribution of the obtained values is shown in Figure S7 along with the results of the Shapiro-Wilk test normality test and the corresponding Q-Q plot. Both tests indicate that the distribution is non-normal, with strong asymmetry and a long one-sided tail. We therefore apply a Box-Cox transformation to such distribution, selecting the transformation parameter $\lambda$ by maximum likelihood optimisation using the `scipy.stats.boxcox` library, which gives $\lambda = -0.202$. The transformed data, shown in Figure S8, satisfy the normality assumptions, allowing application of the $3\sigma$ criterion to detect significant outliers. As shown in Figure S8, no outlier countries are identified. This confirms that sample sizes are consistent across indicators and countries; consequently, this criterion does not modify the set of retained countries.

#### S3.2 Facebook coverage

We assess the representativeness of the UMD Global CTIS and Delphi US CTIS datasets, which are based on surveys administered through Facebook. Although this data-collection strategy is innovative and potentially broad in coverage [11], it raises questions regarding sample representativeness, because Facebook usage is not uniform across countries owing to cultural factors, digital divides, and political or regulatory constraints [12, 13, 14]. In countries where Facebook coverage is low, survey responses may not adequately represent the broader population. Hence, we evaluate Facebook coverage rates, to exclude countries with a high risk of systematic sampling bias. We acknowledge that Facebook statistics may be affected by inactive accounts and by the migration of younger users toward other social media platforms; however, reports from Statista and DataReportal indicate that, in 2021 and 2022, Facebook was still the main social network used globally, with little demographic differences. We therefore use Facebook coverage as a proxy to identify potential systematic biases in access to the online surveys.

To perform this assessment, we use NapoleonCat (https://napoleoncat.com/stats/), which provides monthly statistics on Facebook usage across countries. The consulted archive includes data for May 2021 and June 2022, corresponding to the endpoints of the analysed period. This analysis is performed only at the country level: due to limited data availability, state-level assessment is not possible, for example for individual US states, and the United States is therefore represented as a single entity in terms of Facebook usage, summarising average national coverage.

The NapoleonCat analysis confirms that Facebook usage varies substantially across countries. Figure S9 shows the percentage of the population using Facebook in the countries retained after the initial filtering step. Between May 2021 and June 2022, Facebook usage increased in most countries. However, in countries such as India and Egypt, coverage remained relatively low even in 2022. These trends qualitatively align with the diffusion of mobile devices in each country, reported by DataReportal (see, e.g., https://datareportal.com/reports/digital-2022-chile and url:https://datareportal.com/reports/digital-2022-india for the two extremes in the chart).

Retaining only countries with Facebook coverage of at least 50% leads to the exclusion of India and Egypt from the dataset.

#### S3.3 Epidemic indicators coverage

The availability of testing data is essential for epidemiological indicators, since reported cases should reflect actual epidemic diffusion rather than insufficient testing coverage. We therefore exclude countries that never reported the total number of tests performed. For the remaining countries, we further verify that testing data were reported with sufficient frequency and continuity. Countries with excessive data gaps or substantial discontinuities in reporting are excluded from the analysis. This assessment relies on the OWID and JHU CSSE datasets, as described in the Main Text.

Figure S10 shows missing values of total test numbers for each country and date, marked in black; Figure S11 analogously shows missing values of total case numbers, marked in black, for each country and date.

Based on Figure S10, Brazil and Ukraine are excluded from the analysis because they reported total testing data only during the initial phase of the analysed period. Conversely, countries such as Germany and Poland are retained because their testing data reports show a regular and consistent pattern. Vietnam is excluded because it began reporting testing data late and provided data only for a short interval. Moreover, Brazil, Ukraine and Vietnam had test positivity rates exceeding 50% for extended periods, suggesting severe under-testing (see Figure S12). Alabama and Venezuela are also removed from the final list of countries, because they never reported total testing data. Based on Figure S11, Taiwan is also excluded because of insufficiently reliable case data.

Using public dashboards such as https://coronavirus.jhu.edu/testing/individual-states, we further observe that most US states have test positivity rates that are very high or unavailable for much of the analysed period. This limitation, together with the unavailability of the CHI indicator at the US state level, leads to the exclusion of US states from the analysed dataset, as discussed below.

##### S3.4 CHI availability

As a final selection criterion, we require availability of the CHI indicator at the spatial resolution used in the analysis, for all the countries to be included in the analysis.

In particular, for US states, the OxCGRT indexes do not include H6, corresponding to facial coverings, at the state level, as reported in the README file of the OxCGRT GitHub repository. This limitation, together with the high (or non-reported) test positivity rates discussed above, leads to the exclusion of US states from the analysed dataset.

#### S4 Univariate analysis

We provide here additional details on the univariate analysis, and on the multivariate analysis performed within individual driver classes, presented in the Main Text, subdivided per driver class.

##### S4.1 Beliefs

Here, we interpret the outliers in the Beliefs boxplot show in Figure 3 of the Main Text. While Malaysia experienced political turmoil and strict mask mandates, which may explain its outlier behaviour [15], a more specific interpretation is needed for Japan, Thailand and New Zealand.

Japan and Thailand are Asian countries where mask use is common in daily life due to cultural practices and social norms [16]; see also https://web-japan.org/trends/11_culture/pop202008_mask-culture.html. For both countries, the Containment and Health Index (CHI) varies substantially over time, while mask wearing remains consistently very high and relatively constant (see Figure S13). This discrepancy suggests that mask-wearing behaviour is driven more by established habits than by government enforcements on the occasion of the pandemic. Consistently, the Shannon entropy of the full time series, which is $H = -1/Z \cdot \sum_{i=0}^{100} p(i) \log p(i) \in [0;1]$, where $p(i)$ is the probability of observing a certain value $i \in \{0, 1, \ldots, 100\}$ and $Z$ is a normalization constant, gives $H^{\text{Japan}}_{\text{mask usage}} = 0.85$ and $H^{\text{Japan}}_{\text{worried COVID}} = 0.91$, indicating a relatively persistent situation subject to random fluctuations. Similarly, for Thailand, $H^{\text{Thai}}_{\text{mask usage}} = 0.87$ and $H^{\text{Thai}}_{\text{worried COVID}} = 0.89$.

New Zealand differs from these cases because it had no established culture of routine mask use. As shown in Figure S14, mask use increased sharply in September 2021, when mask mandates were very stringent, and then gradually decreased. The increase in September 2021 therefore appears to be primarily associated with imposed requirements rather than personal risk perception. Consistently, New Zealand is no longer an outlier when analysing the correlation between mask wearing and CHI. The observation that the increase in mask use is predominantly driven by government restrictions is corroborated by the fact that, in April 2022, mask use decreased despite increasing cases and deaths (*cf.* Figure S14), possibly due to pandemic fatigue.

##### S4.2 Awareness

Similar considerations help explain the outliers in the correlations between mask-wearing behaviour and awareness indicators (see Figure 3 of the Main Text).

A separate issue concerns the broad distributions observed for Awa5 ("received news from politicians") and Awa6 ("received news from journalists"). These two indicators display substantially higher variability across countries than the other awareness indicators. To investigate this heterogeneity, we analyse the distributions of country-level correlations between mask-wearing and Awa5 (respectively,

Awa6) in Figure S15 (respectively, in Figure S16). The dashed vertical lines mark the boundaries between three clusters corresponding to low, intermediate, and high correlation values, identified using $K$-Means clustering with $K = 3$. The values for each country are additionally coloured by geographical area (Europe, North America, Oceania, Latin America, Asia) to assess possible geographical patterns. In Figure S15, which considers information received from politicians, Asian countries occupy the lower end of the correlation distribution, European countries occupy the upper end, and Latin American countries are mainly located in the intermediate range. In Figure S16, considering information received from journalists, Asian and Latin American countries are concentrated toward the lower end of the distribution, whereas European countries occupy the central and upper ranges. These patterns suggest that, during the COVID-19 pandemic, information received from journalists and politicians was more strongly associated with mask-wearing behaviour in European countries. This was not observed in Asian countries, where correlations were weak or even negative. Such differences may reflect heterogeneous communication strategies during the pandemic, different media ecosystems, or different levels of trust in political and journalistic institutions. Overall, the association between behavioural responses and information received from politicians or journalists varies markedly across geographical regions; this observation supports the use of geographically tailored behavioural-epidemiological models incorporating awareness, and highlights the importance of accounting for the effectiveness of narratives and social actors in pandemic communication.

#### S4.3 Mutual relationships among drivers

To further justify the use of a multivariate and nonlinear Random Forest approach, we assess whether different classes of drivers exhibit mutual relationships. This analysis helps identify multicollinearity patterns that would hinder the applicability of simpler MLR models including all drivers simultaneously, and highlights correlations and interdependencies among the drivers that may inform more realistic behavioural-epidemiological models, as well as more effective decision-making and interventions.

Figure S17 shows pairwise correlations between selected classes of drivers. Panels a-d show that the **CHI** correlates substantially with **beliefs** and **awareness**, supporting previous longitudinal studies about beliefs about efficacy concurring in adherence to policy mandates [16]; on the contrary, it correlates less strongly with **trust** and **epidemiological indicators**. Granger causality analysis indicates that CHI Granger-causes both belief indicators, but only the belief in mask effectiveness significantly Granger-causes the CHI. These observations are consistent with the interpretation that, while public beliefs are shaped by institutional decisions, government decisions may be also influenced by knowledge and perception of the effectiveness of preventive measures. However, although government interventions do influence public beliefs, this effect is limited quantitatively: MLR performed separately for each belief gives $R^2 = 0.23$ for fear of infection and $R^2 = 0.13$ for belief in mask effectiveness. Thus, governmental interventions have only a modest direct influence on public beliefs. These are average results, and the long whiskers in the boxplots (panel a) indicate substantial country-level variability.

Regarding the relationship of **CHI** with **awareness** (panel b), increases in the Containment Health Index are generally associated with increases in the percentage of individuals receiving information from specific sources. The main exception is awareness through journalists (Awa6), whose distribution is broad and includes countries with negative correlations. Figure S18 shows this distribution in more detail, with countries clustered as described above. A clear geographical subdivision emerges: European countries show moderately high correlations, while Asian countries and most Latin American countries show low to negative correlations. In countries with positive correlations, such as those in Europe, receiving information from journalists appears to be associated with stronger containment measures. Conversely, in countries with negative correlations, such as many Asian countries, information dynamics follow trends distinct from government interventions. These differences might reflect cultural practices in information consumption, the presence of alternative information sources, constraints on press freedom, or disinformation processes [17, 18, 19]; testing these mechanisms is left to future work. Moreover, the Granger causality analysis suggests that CHI Granger-causes awareness through all information sources, while awareness through experts, WHO and government health authorities precede changes in CHI, reflecting that government decisions were largely influenced by local and global experts and health authorities.

Correlations and mutual causal relationships between the **CHI** and **trust** (panel c) are negligible and show high variability, and the Granger causality test only indicates unidirectional relationships between CHI and trust indicators. Overall, trust does not appear to have a relevant influence on CHI.

By contrast, although correlations between the **CHI** and **epidemiological indicators** are low (panel d), Granger causality analysis indicates that incidence levels Granger-cause the CHI, and vice versa, as expected. Instead, only the CHI significantly Granger-causes changes in new deaths, whereas the opposite relationship is not significant. This is consistent with the fact that the adopted non-pharmaceutical interventions were effective in reducing casualties, while most governmental decisions were taken in response to increases in daily cases, before deaths accumulated [20, 21].

Figure S17e shows the correlation between **New Cases** (incidence, not mediated by news sources) and **beliefs**. The two boxplots display different trends. The correlation between new cases and fear of infection (Bel1) is weak but positive, with a median around 0.2, consistently with the hypothesis that increasing cases may increase fear. Conversely, the correlation between new cases and belief in mask effectiveness (Bel2) has a median around −0.1; a plausible hypothesis is that increasing infections may be associated with reduced confidence in mask effectiveness. In both cases, the whiskers of the boxplots are very broad, indicating high variability across countries. To test these hypothesis, we performed two Granger causality tests. Fear of contracting COVID-19 Granger-causes new cases, suggesting that changes in infection dynamics may be preceded by changes in fear levels, consistent with the patterns observed in the distribution of correlations. For belief in mask effectiveness, the Granger test result is not statistically significant ($p = 0.0537$), indicating that the hypothesis is not supported at the chosen significance threshold. In both cases, however, MLR yields very low $R^2$ values. These results suggest that epidemiological indicators alone have limited behavioural relevance for the general public, either because individuals do not consult epidemiological dashboards regularly or because such information affects behaviour primarily when mediated by information sources.

Consistently, Figure S17f and Figure S17g show substantially stronger correlations between **beliefs** and **awareness** indicators, except for journalists, as discussed above. The numerical values of incidence or deaths therefore appear to carry limited behavioural meaning on their own. The nonlinear relationships and mediating effects between beliefs and epidemiological indicators should be investigated in future work to support the development of more accurate behavioural-epidemiological models.

Finally, we confirm that **trust** is not a significant driver in this analysis (see Figure S17c for its correlation with the CHI). We also tested whether weighting awareness from each source by trust in that source improves the association with mask-wearing behaviour, constructing the composite indicator $\text{Awa}_k/\text{Trust}_k$ for each source $k$. As shown in Figure S18b, the resulting correlations are even weaker than those obtained using awareness alone (shown in Figure 3 of the Main Text), confirming that trust plays a negligible role in shaping mask-wearing behaviour and the other drivers considered here.

Table S1: Indicators of OxCGRT; adapted from [2].

| Policy measure | ID | Name |
|---|---|---|
| **Containment and Closure Policies** | | |
| | C1 | School closure |
| | C2 | Workplace closing |
| | C3 | Cancel public events |
| | C4 | Restrictions on gatherings |
| | C5 | Public Transportation |
| | C6 | Stay at home order |
| | C7 | Restrictions on internal movement |
| | C8 | International travel controls |
| **Health System Policies** | | |
| | H1 | Public information campaigns |
| | H2 | Testing policy |
| | H3 | Contact tracing |
| | H4 | Emergency investment in healthcare |
| | H5 | Investment in vaccines |
| | H6 | Facial coverings |
| | H7 | Vaccination policy |
| | H8 | Protection of elderly people |
| **Economic Policies** | | |
| | E1 | Income support |
| | E2 | Debt/contract relief for households |
| | E3 | Fiscal measures |
| | E4 | Providing Support to other countries |
| **Vaccination Policies** | | |
| | V1 | Vaccine prioritisation |
| | V2 | Vaccine eligibility/availability |
| | V3 | Vaccine financial support |
| | V4 | Mandatory vaccination |
| **Miscellaneous Policies** | | |
| | M1 | Other responses |

Table S2: OxCGRT summary indicators with the related specific components; adapted from [2].

| **Index** | C1 | C2 | C3 | C4 | C5 | C6 | C7 | C8 | E1 | E2 | E3 | E4 | H1 | H2 | H3 | H4 | H5 | H6 | H7 | H8 | M1 | V1 | V2 | V3 | V4 |
|---|---|---|---|---|---|---|---|---|---|---|---|---|---|---|---|---|---|---|---|---|---|---|---|---|---|
| Government response index | X | X | X | X | X | X | X | X | X | X | | | X | X | X | | | X | X | X | | | | | |
| Containment and health index | X | X | X | X | X | X | X | X | | | | | X | X | X | | | X | X | X | | | | | |
| Stringency index | X | X | X | X | X | X | X | X | | | | | X | | | | | | | | | | | | |
| Economic support index | | | | | | | | | X | X | | | | | | | | | | | | | | | |
| Legacy stringency index | X | X | | | X | | | X | | | | | X | | | | | | | | | | | | |

Table S3: Face Covering Index Scale; adapted from [2].

| Value | Policy Description |
|---|---|
| 0 | No policy |
| 1 | Recommended |
| 2 | Required in some specified shared/public spaces outside the home with other people present, or some situations when social distancing not possible |
| 3 | Required in all shared/public spaces outside the home with other people present or all situations when social distancing not possible |
| 4 | Required outside the home at all times regardless of location or presence of other people |

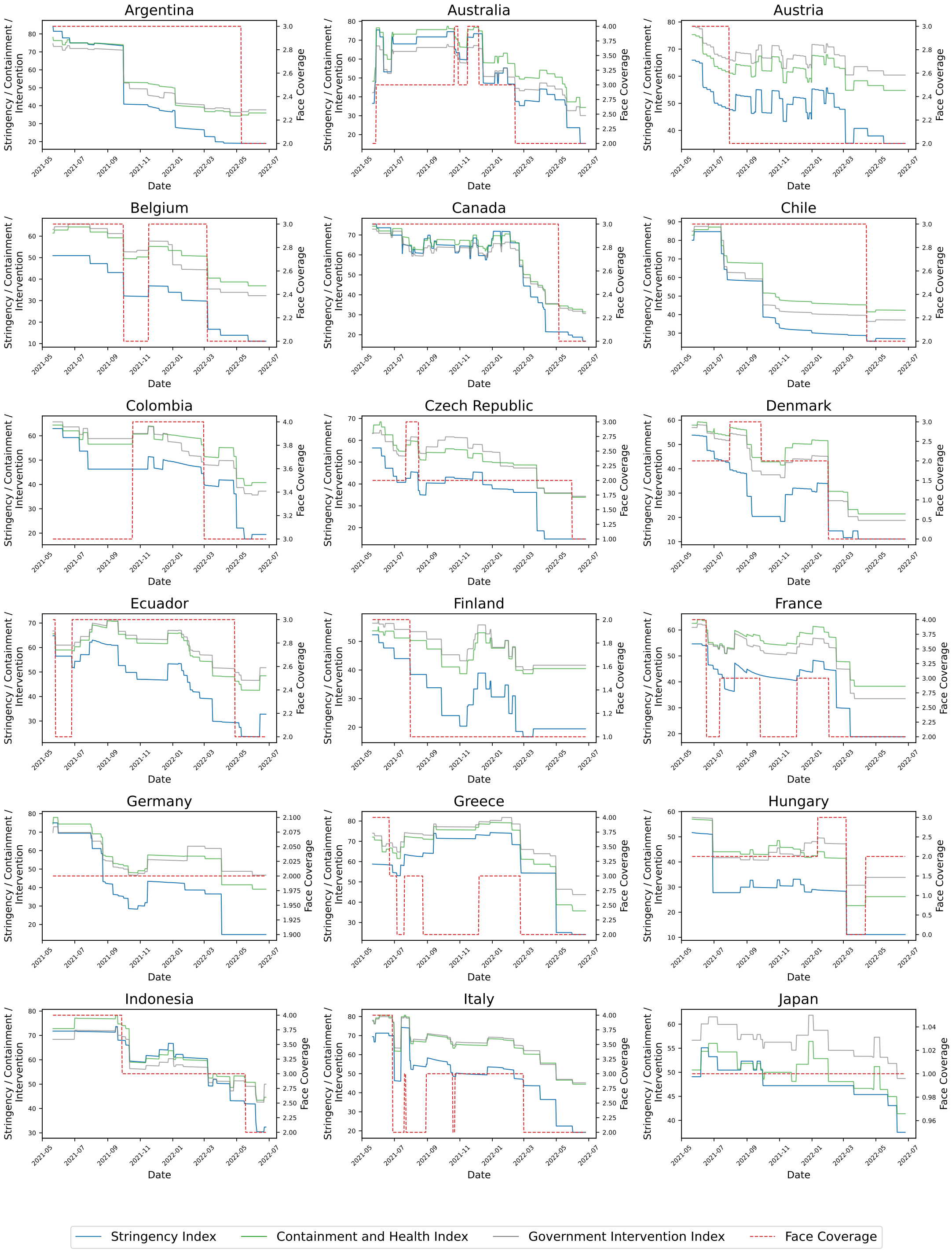


Figure S1: Comparison over time between Stringency Index, Containment and Health Index, Government Intervention Index and Face Covering indicator (Part I).

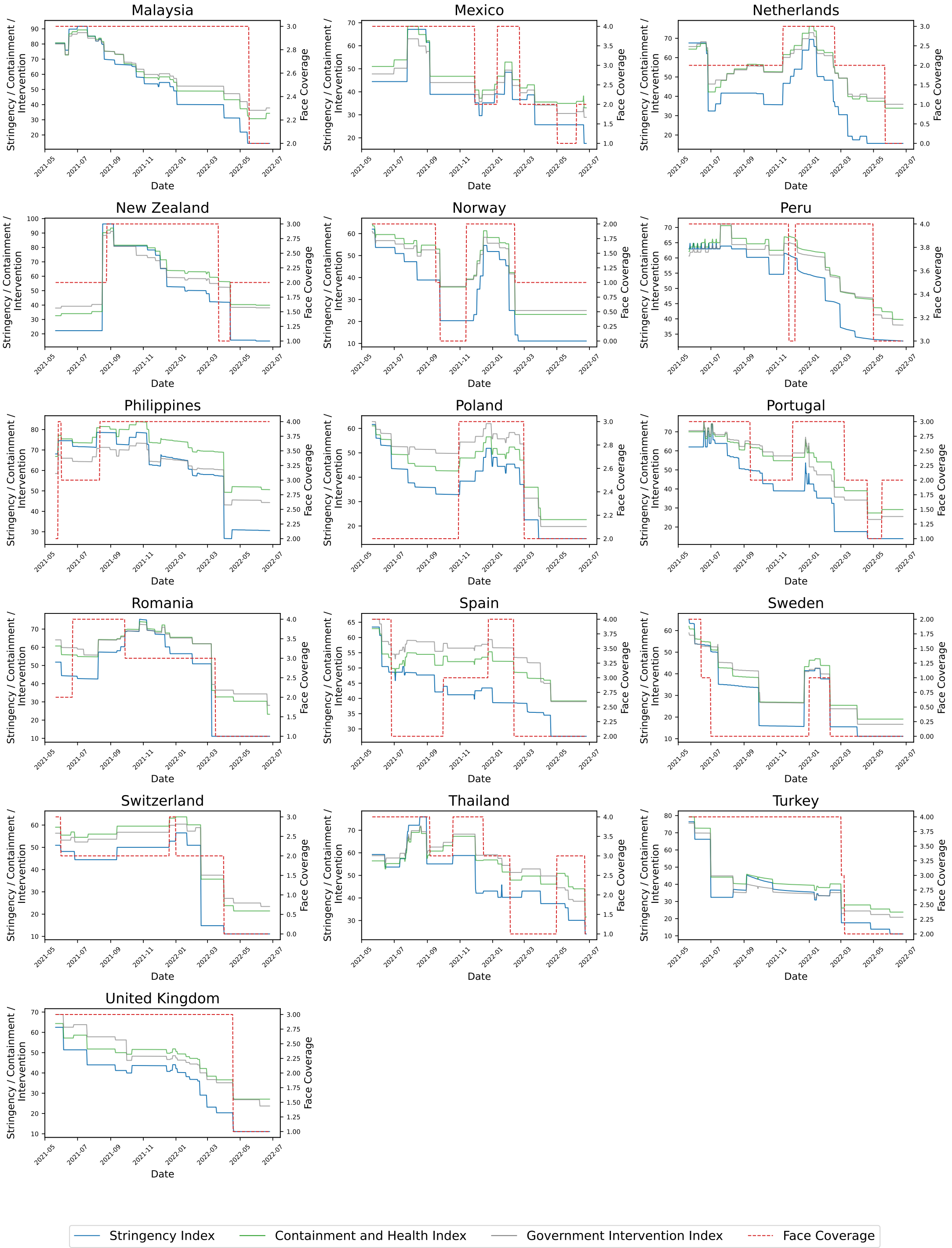


Figure S2: Comparison over time between Stringency Index, Containment and Health Index, Government Intervention Index and Face Covering indicator (Part II).

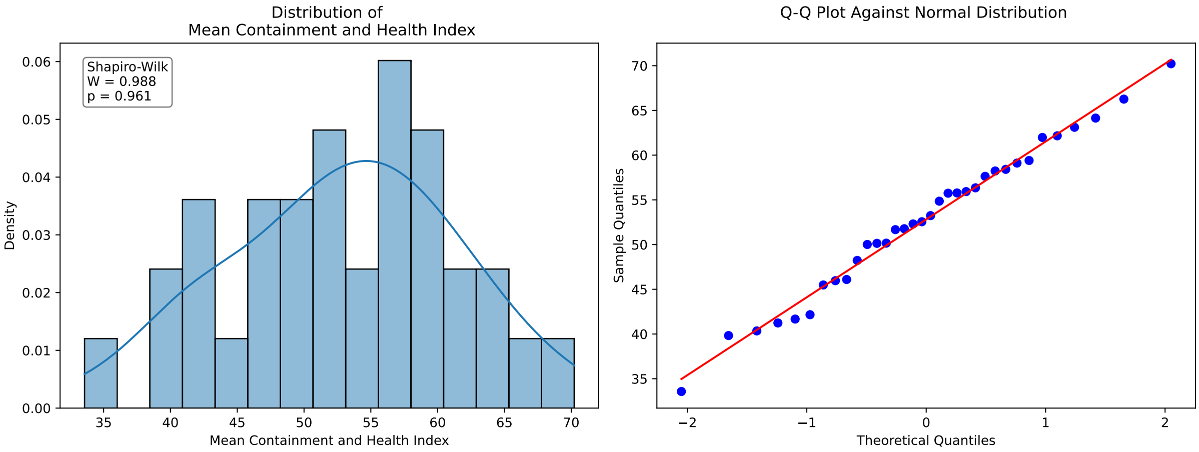


Figure S3: Distribution of the average Containment and Health Index across countries, together with the outcome of the Shapiro-Wilk normality test and the corresponding Q-Q plot.

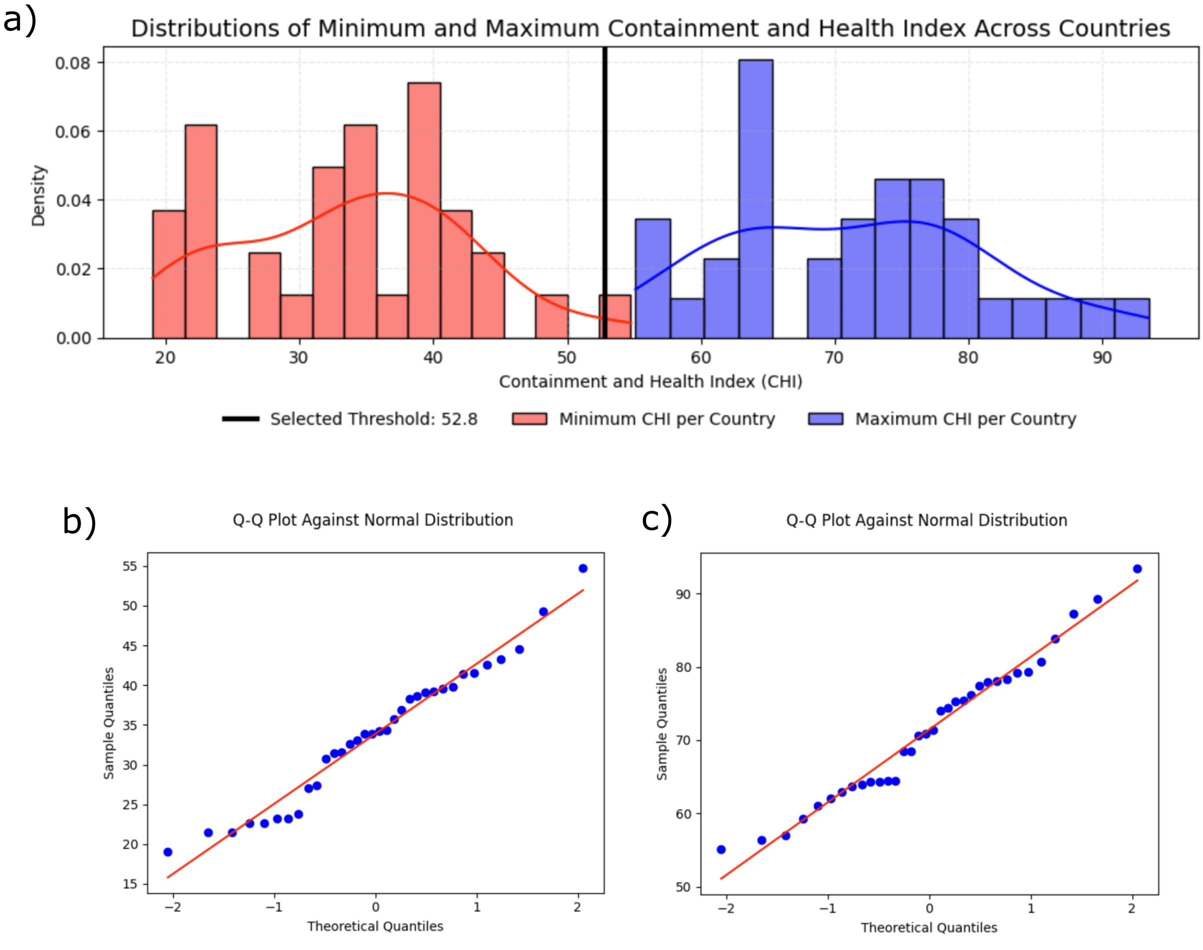


Figure S4: a) Distributions of the maximum (red) and of the minimum (blue) values of Containment and Health Index across countries. For visual comparison, the vertical line represents the threshold identified from the mean CHI values: all maxima fall above it, and almost all minima are below it. b) and c) The Q-Q plots corresponding to distributions of minima and maxima, respectively. The outcome of the Saphiro-Wilk test for the distribution of minima is: $W = 0.9663$, $p = 0.3674$; for the distribution of maxima: $W = 0.9702$, $p = 0.4674$. In both cases, we cannot discard the null hypothesis that the distributions are normal.

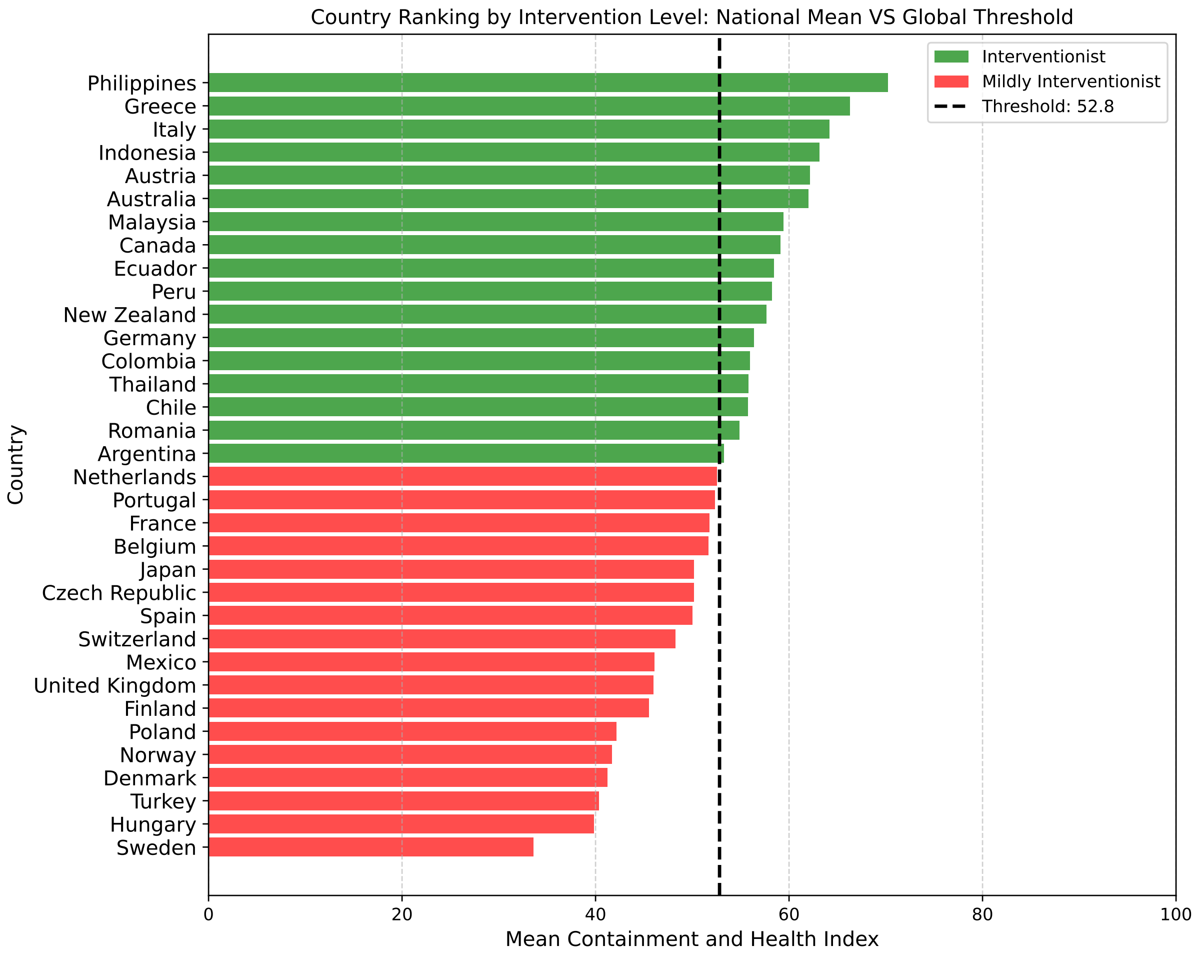


Figure S5: Ranking of countries according to the average value of the Containment and Health Index over time. The vertical dashed line (Threshold) corresponds to the mean of the distribution across countries and serves as a visual reference distinguishing more and less interventionist countries.

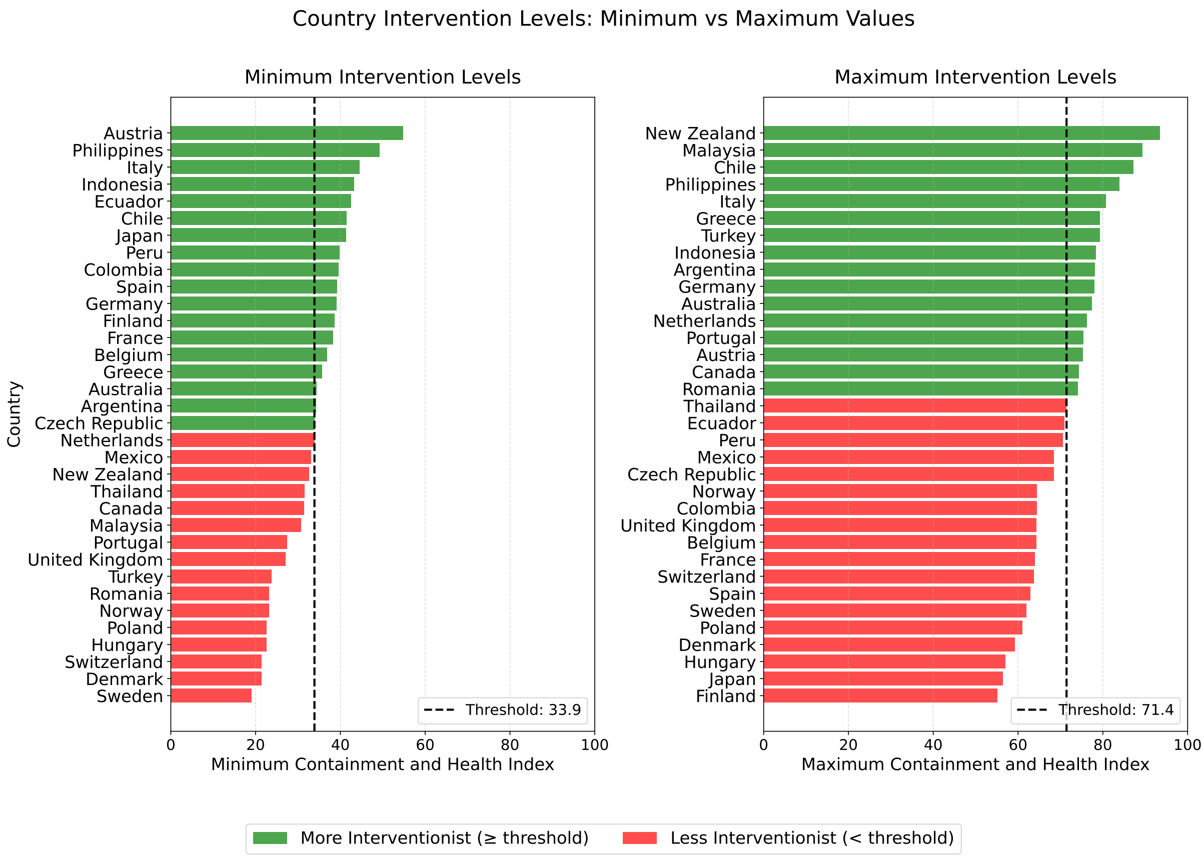


Figure S6: Ranking of countries according to the minimum (left) and the maximum (right) values of the Containment and Health Index over time. The vertical lines (Threshold) represent the corresponding mean values of the two distributions across countries and serve as visual references distinguishing more and less interventionist countries. The rankings of countries based on the values of the minimum, the maximum and the mean (Figure S5) CHI are all different, reflecting the uneven adoption of stringency measures by the considered countries.

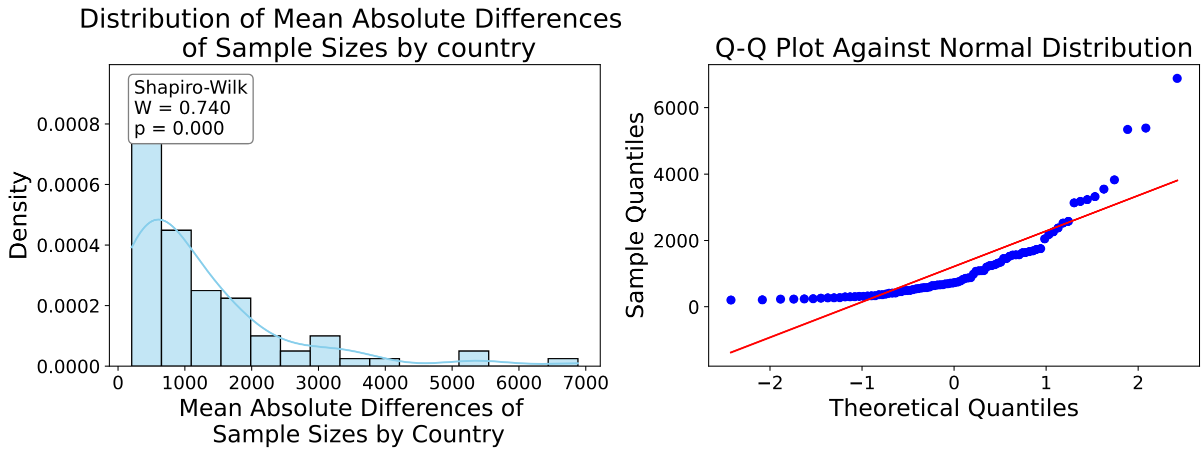


Figure S7: Distribution of the mean absolute differences in sample sizes, see (S3), across countries, together with the outcome of the Shapiro-Wilk normality test and the corresponding Q-Q plot.

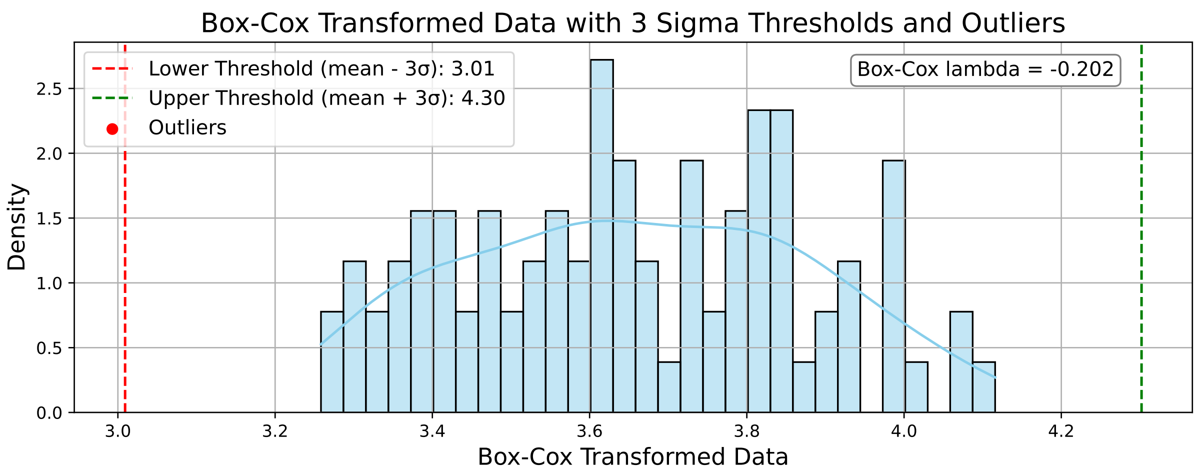


Figure S8: Distribution of the Box-Cox transformed data and identification of potential outliers (there are none).

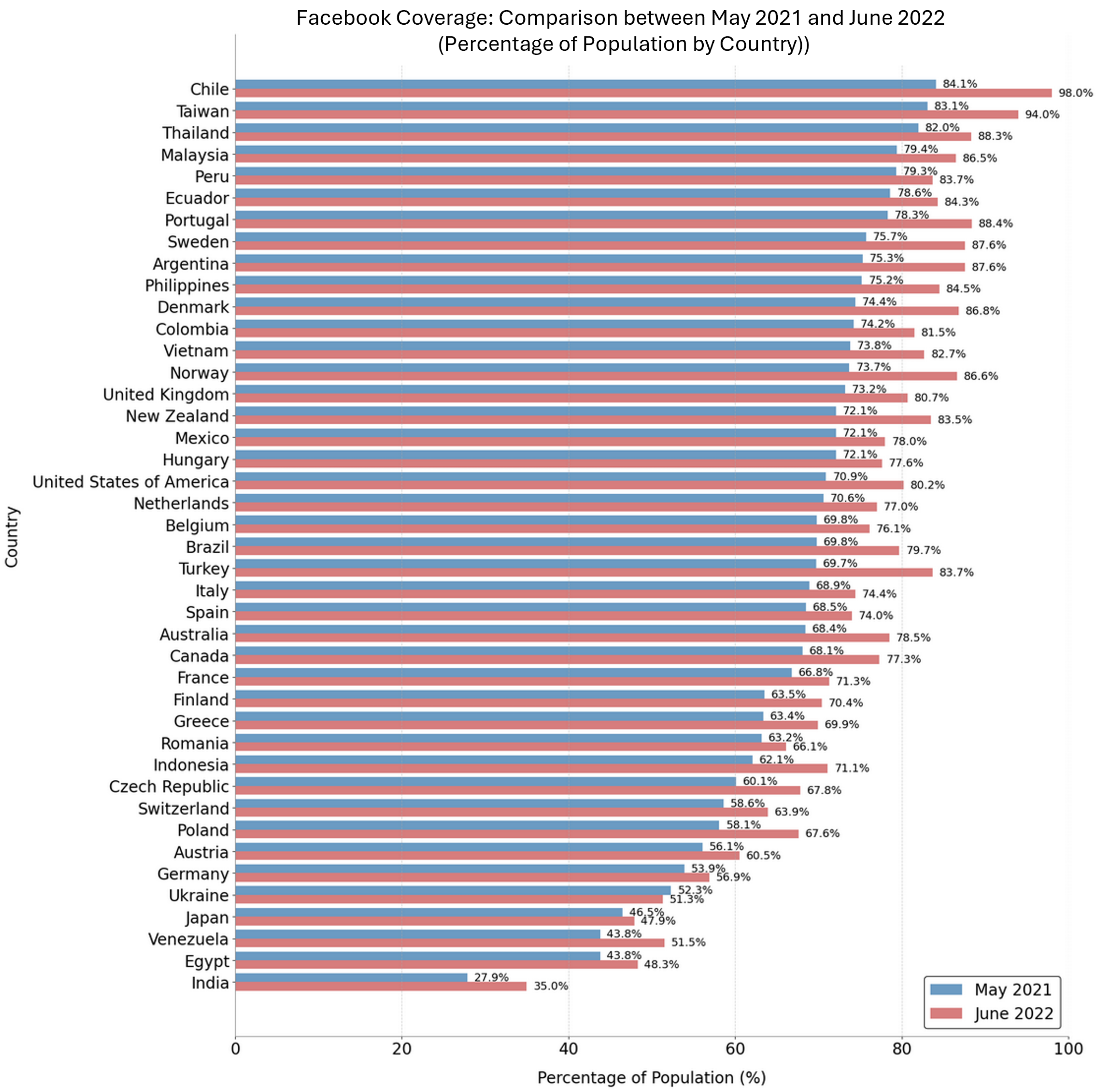


Figure S9: Facebook coverage in May 2021 and June 2022 for all countries retained after the first filtering step.

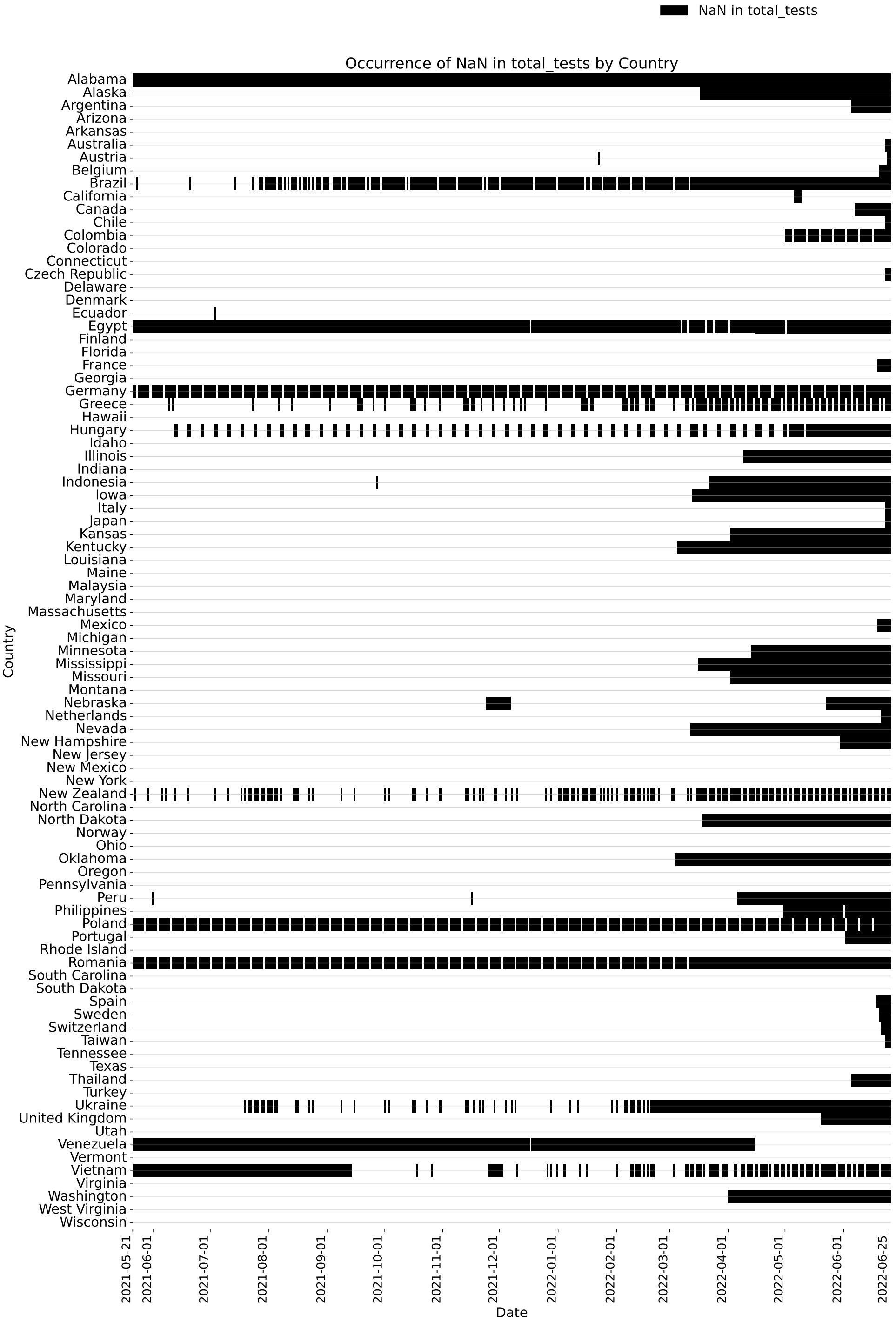


Figure S10: Presence and absence of total testing data in the OWID and JHU CSSE datasets. Missing values (NaN) are shown in black.

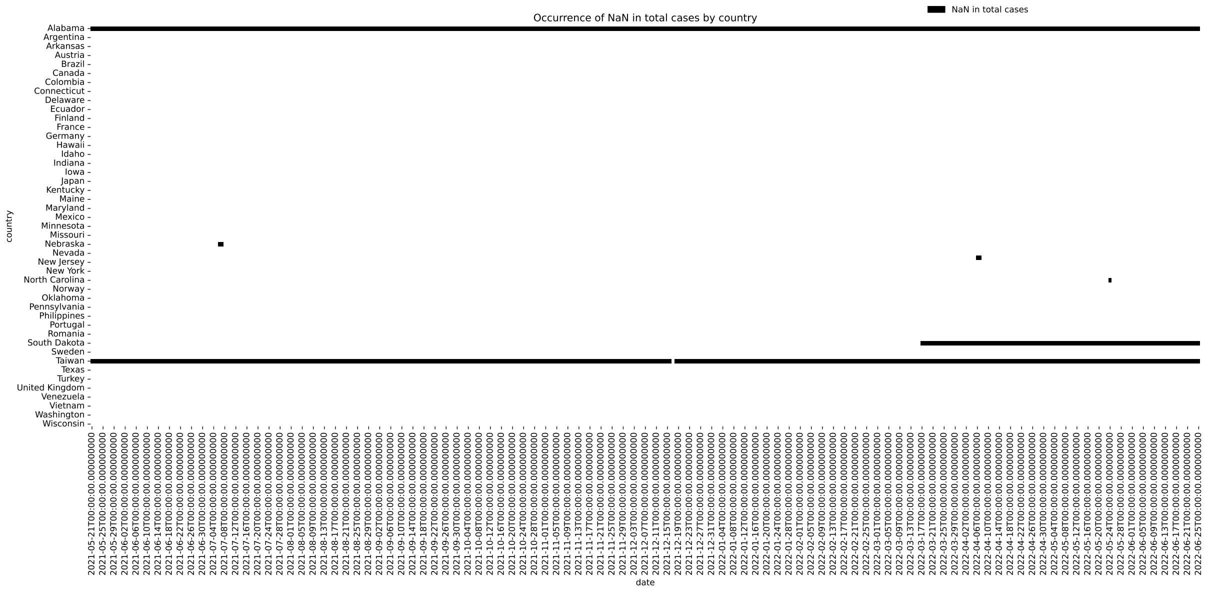


Figure S11: Presence and absence of total case data in the OWID and JHU CSSE datasets. Missing values (NaN) are shown in black.

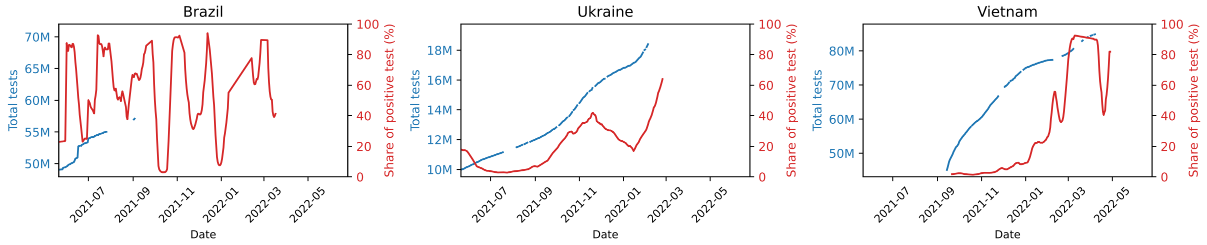


Figure S12: Time evolution of total testing counts and test positivity rates for Brazil, Ukraine, and Vietnam.

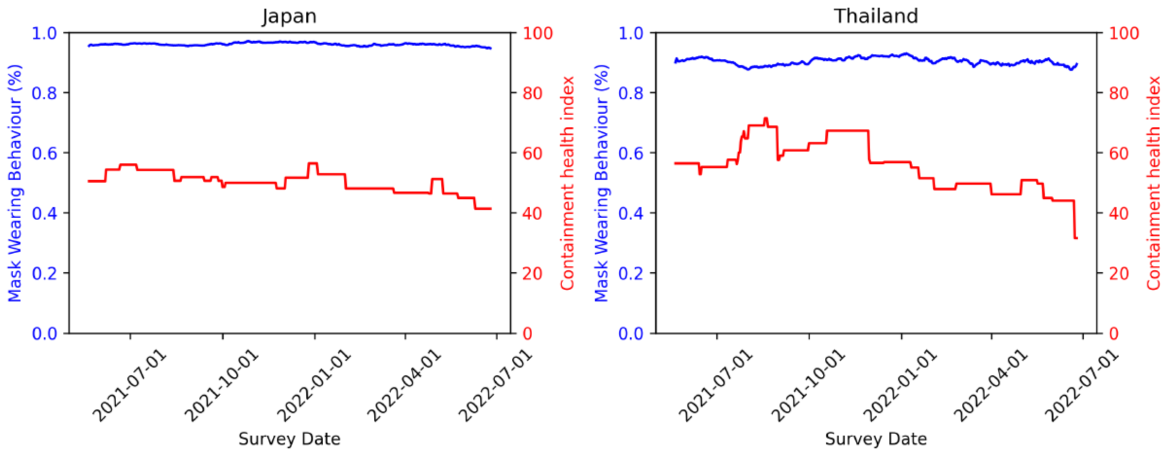


Figure S13: Time evolution of mask-wearing behaviour and of the Containment and Health Index in Japan and Thailand.

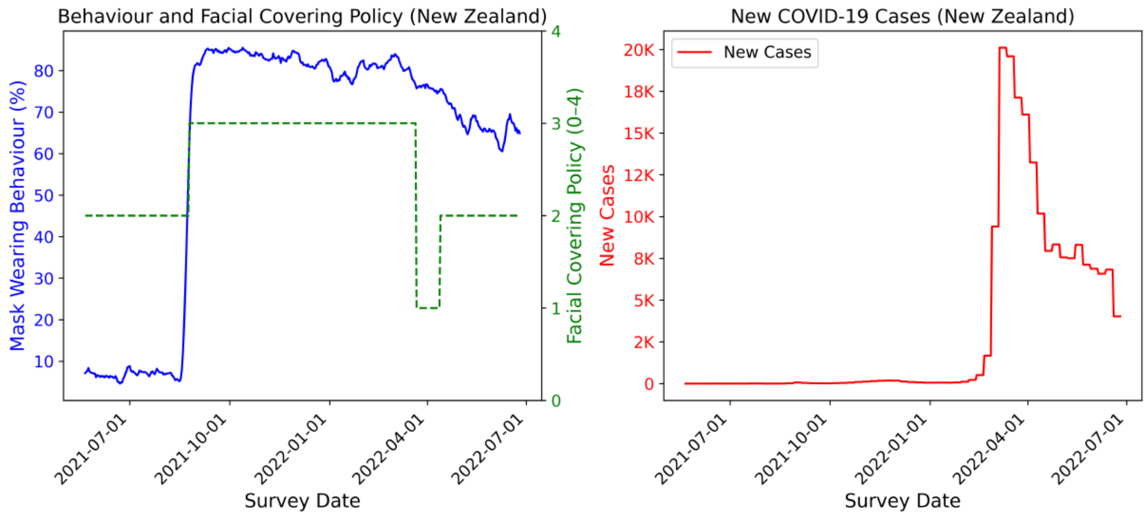


Figure S14: Time evolution of mask-wearing behaviour, facial-covering policies, and new COVID-19 cases in New Zealand.

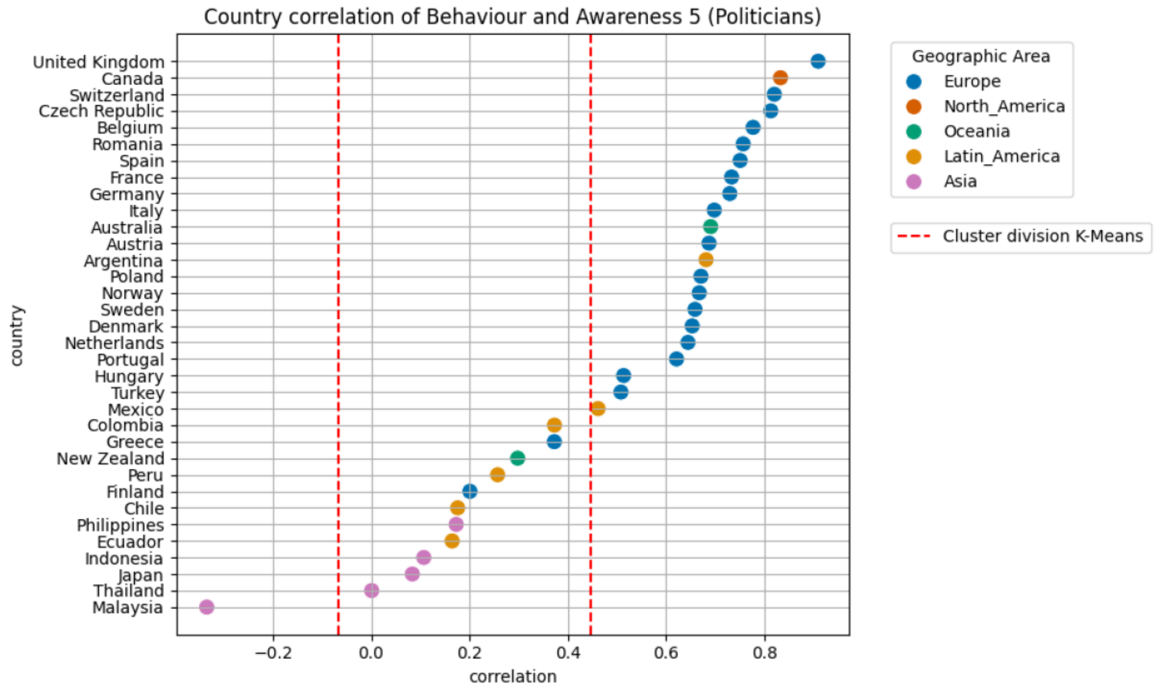


Figure S15: Country-level correlations between mask-wearing behaviour and Awa5 ("received news from politicians"), showing the three clusters identified through $K$-Means (separated by dashed thresholds). Countries are coloured according to geographical area.

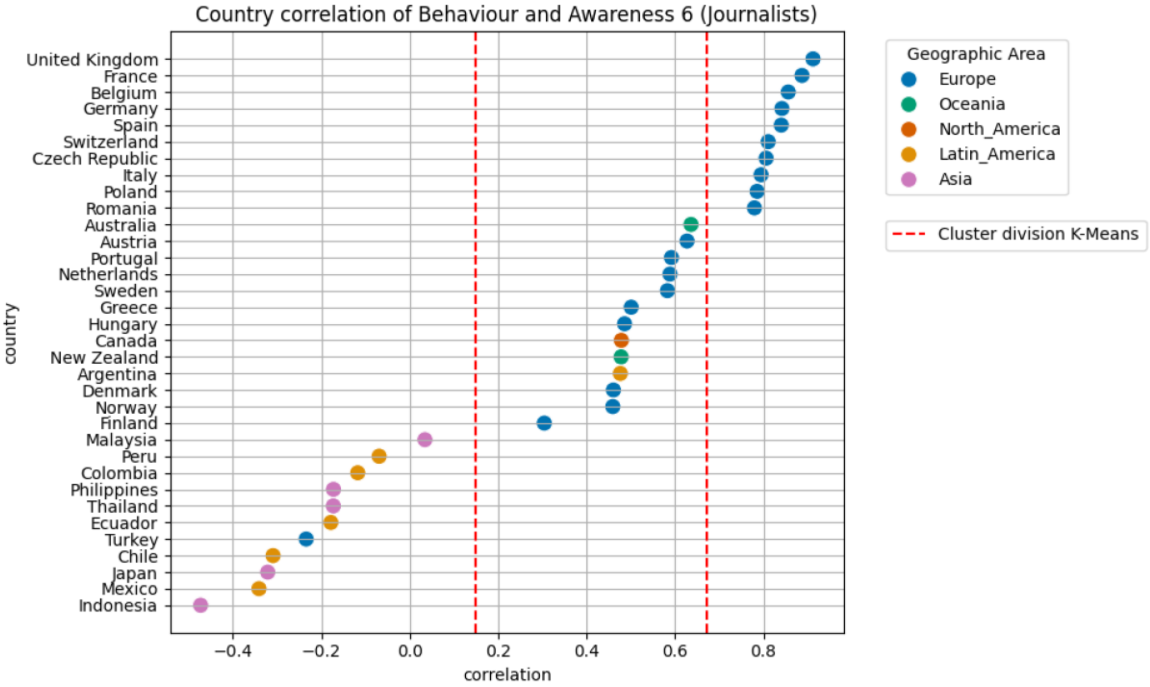


Figure S16: Country-level correlations between mask-wearing behaviour and Awa6 ("received news from journalists"), showing the three clusters identified through $K$-Means (separated by dashed thresholds). Countries are coloured according to geographical area.

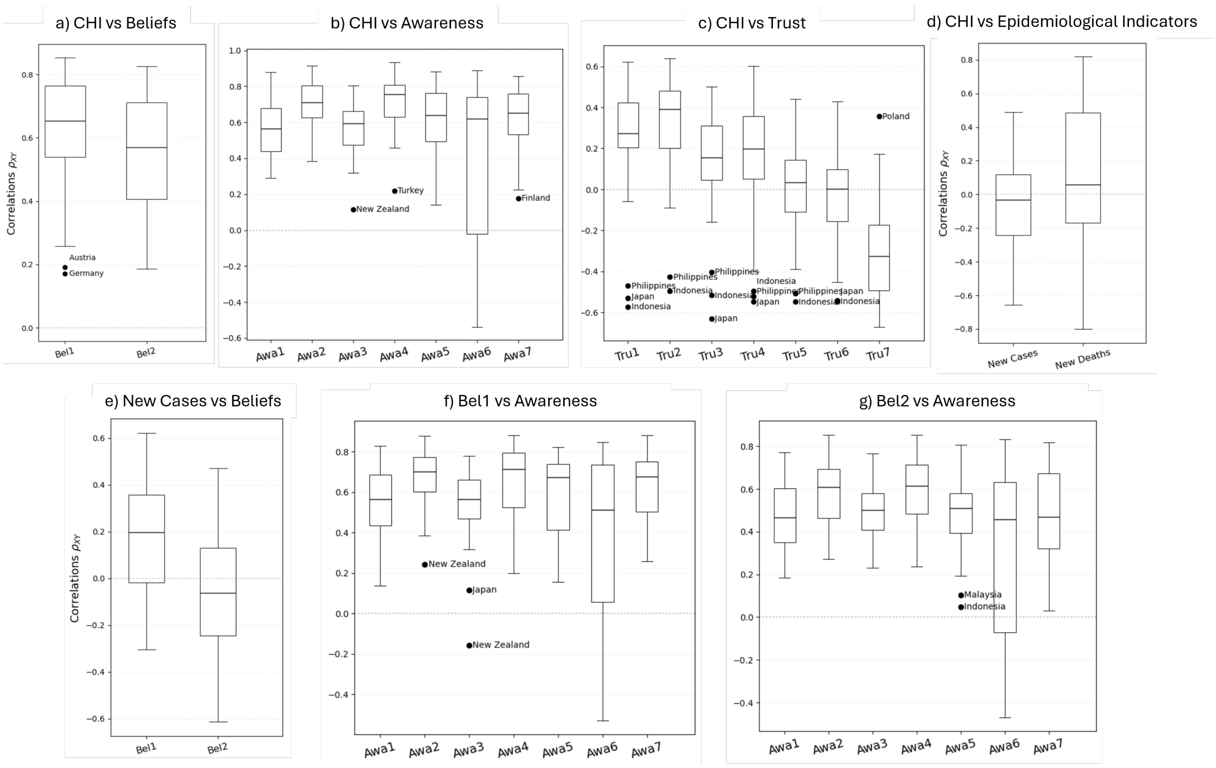


Figure S17: Pairwise correlations between drivers in different classes. Belief-related indicators are fear of COVID-19 infection (Bel1) and perceived effectiveness of mask wearing (Bel2). Awareness and trust refer respectively to local health workers (Awa1, Tru1), experts (Awa2, Tru2), WHO (Awa3, Tru3), government health authorities (Awa4, Tru4), politicians (Awa5, Tru5), journalists (Awa6, Tru6), friends and family (Awa7, Tru7). Panels a-d: correlations between the CHI and drivers in the other classes (beliefs, awareness, trust and epidemiological indicators, respectively). Panel e: correlations between new cases and belief-related indicators. Panels f-g: correlation between beliefs and awareness indicators.

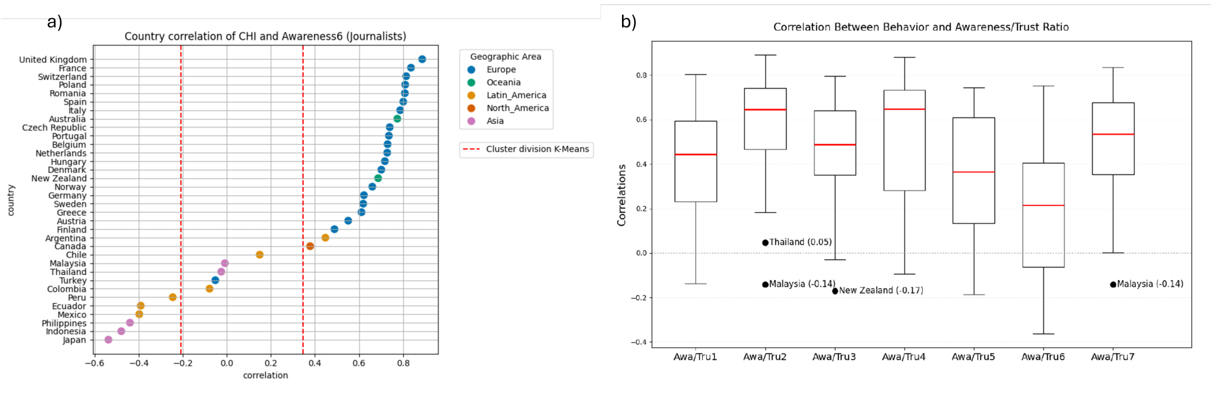


Figure S18: a) Country-level correlations between the Containment and Health Index and Awa6 (“received news from journalists”), showing the three clusters identified through $K$-Means (separated by dashed thresholds). Countries are coloured according to geographical area. b) Boxplots of correlations between mask-wearing behaviour and the awareness/trust ratio across countries.
